# Factors associated with vaccination against Seasonal Influenza and SARS-CoV-2 in the UK among FluSurvey participants, 2023-2025

**DOI:** 10.64898/2026.08.25.26361301

**Authors:** L Adams, CH Watson, G Dabrera, RE Green

## Abstract

Seasonal Influenza and COVID-19 vaccination programmes are critical for reducing morbidity and mortality in older adults, yet uptake remains uneven across populations. We aimed to profile vaccination attitudes and examine predictors of COVID-19/influenza vaccination uptake among a UK participatory surveillance system - FluSurvey.

We analysed FluSurvey data from participants aged ≥65 years who were eligible for both vaccines in the 2023–2024 and 2024–2025 Autumn – Winter seasonal campaigns. Descriptive analyses examined self-reported attitudes to influenza vaccination. Logistic regression examined factors (age, sex, socioeconomic status, education, employment, transport, smoking and chronic conditions) associated with influenza and COVID-19 vaccination uptake in each season, adjusting for confounders.

Belonging to a risk group and reducing risk of influenza were frequently reported motivations for influenza vaccination, while building natural immunity and concerns around safety and adverse effects were frequently reported barriers. Individuals vaccinated against COVID-19 were more likely to receive an influenza vaccination (aOR_2023-2024_=13.90 [9.28-21.17]; aOR_2024-2025_=8.54 [5.82-12.60]), and vice-versa (aOR_2023-2024_=13.91 [9.30-21.19]; aOR_2024- 2025_=8.52 [5.81-12.58]). Lower educational attainment was associated with lower odds of COVID-19 vaccination (aOR_2023-2024_=0.59 [0.45-0.78], aOR_2024-2025_: 0.56 [0.39-0.79]). Other results were weaker or demonstrated variation by season.

Our findings highlight recent attitudes and barriers to influenza and COVID-19 vaccination among the FluSurvey cohort, which may inform approaches to improve vaccination coverage in the population.

**Summary:** We aimed to describe influenza and COVID-19 vaccination attitudes and identify factors associated with vaccine uptake among adults aged 65 years and older participating in FluSurvey during the 2023–2024 and 2024–2025 UK autumn–winter seasons. We analysed self-reported vaccination and demographic, socioeconomic, health and lifestyle data from the FluSurvey profile questionnaire, using descriptive analyses to examine reasons for influenza vaccination decisions and multivariable logistic regression to assess predictors of influenza and COVID-19 vaccination uptake. Influenza vaccination uptake was 81% in both seasons, while COVID-19 vaccination increased from 73% to 83%. The most frequently reported motivations for influenza vaccination were belonging to a risk group and reducing influenza risk; common barriers were beliefs in natural immunity and concerns about vaccine safety or adverse effects. Prior vaccination against the other disease was the strongest and most consistent predictor: prior COVID-19 vaccination was associated with influenza vaccination (aOR 13.90, 95% CI 9.28–21.17, in 2023–2024; 8.54, 5.82–12.60, in 2024– 2025), with similar associations in the reverse direction. Lower educational attainment was associated with reduced COVID-19 vaccination. Overall, findings support integrated vaccination approaches and communication addressing perceived risk and safety concerns, while highlighting the need for larger, more representative studies.

## 1. Introduction

Seasonal influenza contributes to morbidity, mortality and subsequent pressures on healthcare services (1). Seasonal influenza vaccination was first introduced in England in the late 1960’s. Since then, the programme has changed to include older adults (65 and above), children and targeted clinical risk groups who are at a higher risk of developing severe illness due to influenza (2). Influenza vaccination is one of the most effective public health interventions for preventing the burden associated with influenza. A meta-analysis of influenza vaccine efficacy in elderly persons estimates vaccine efficacy to be 56% (95% CI, 39% to 68%) for preventing respiratory illness, 53% (CI, 35% to 66%) for preventing pneumonia, 50% (CI, 28% to 65%) for preventing hospitalization, and 68% (CI, 56% to 76%) for preventing death (3).

COVID-19 vaccination is also offered in the autumn to those who are higher risk of severe illness. A cumulative meta-analysis found that the COVID-19 vaccines were effective in preventing severe acute respiratory syndrome coronavirus 2 (SARS-CoV-2) infection (OR = 0.38, [0.23–0.65]) and in reducing the number of COVID-19-related deaths (OR = 0.16, [0.10–0.25]) in elderly people (4).

Despite the benefits of influenza and COVID-19 vaccination, uptake in the UK remains below the World Health Organisation target of 75% coverage among several key groups (5). Recent national data show that while uptake among older adults (aged ≥65 years) is relatively high, 74.9% in the 2024-2025 season for influenza vaccination, coverage in the same group for COVID-19 vaccination is only 59.3% (6). Furthermore, substantial variation exists by demographic and social factors; for example, uptake rates are consistently lower in more socioeconomically deprived areas and among certain population subgroups, and regional differences persist across England (7). This heterogeneity in uptake presents a significant public health challenge, as suboptimal coverage undermines population protection and may contribute to preventable morbidity and health system strain during influenza seasons.

Previous literature suggests that influenza and COVID-19 vaccine uptake is shaped by a complex interplay of factors and varies over time. In the UK, analysis of the General Practice Research Database (GPRD) spanning the 2009/2010 pandemic season found that within clinical risk groups, uptake varied by age and comorbidity burden (8). More recently a nationwide cohort study investigated COVID vaccine hesitancy from 2020 to 2022. They reported that 3.3% of participants presented some sort of hesitancy across the study period and this peaked at 8.0% in early 2021 and reduced to 1.1% in early 2022. However, these results are likely to be influenced by the COVID-19 pandemic, only present reasons against vaccination and are limited to COVID-19 (9). Meanwhile, a systematic review of studies among older adults (≥ 65 years) identified a broad array of structural, intermediate, and system-level determinants influencing seasonal influenza vaccine acceptance; including socioeconomic status, education, ethnicity, living situation, health status, perceived health and susceptibility, prior vaccine experience, and trust in providers or public health messaging (10). However, the review is now dated (search up to 2011), may not reflect contemporary UK health-service delivery (e.g. expanded vaccination settings), and its findings - drawn from heterogeneous international studies - may not accurately characterise the UK context.

FluSurvey is a web-based participatory surveillance system used to monitor trends in influenza and other respiratory viruses among the UK population. In addition to weekly symptom surveys, participants complete a background “profile” questionnaire on demographics, vaccination (influenza, COVID-19, RSV) and risk factors of respiratory illnesses. The profile survey may be updated at any time, such as following vaccination, and participants are additionally asked follow up questions as to reasons why they received or did not receive an influenza vaccine. This presents a unique opportunity to understand both vaccination behaviours and predictors of vaccination uptake among a well-characterised cohort following the COVID-19 pandemic.

This study will describe FluSurvey participants and the equivalent group from the English population. Reasons for vaccination choices will be explored, and logistic regression models will be used to identify factors associated with vaccination uptake.

## 2. Methods

Individuals aged 18 and over who reside in the UK are eligible to register and participate in FluSurvey (11). Registered participants may additionally report on behalf of other household members including children. The analysis will include the FluSurvey cohort who are 65 and over from the 2023-2024 (week 44 2023 to week 16 2024) and 2024-2025 (week 47 2024 to week 14 2025) seasons, ensuring all participants involved in the analysis were eligible for influenza and COVID-19 vaccination during these time periods. In all analyses, profile questionnaires (submitted at the start of each season and regularly prompted to be updated throughout the season) will be used to derive variables of interest. Profile questionnaires will be deduplicated to retain one per participant in each season. The first one where vaccination is reported will be used for those vaccinated and the first one of the season for those who never report a vaccination.

### Variables

COVID-19 and influenza vaccination status within the season will be defined individually as binary variables (yes/no) based on self-reported responses from the profile survey. Demographic characteristics (age, sex, and ethnicity), geographic deprivation (Index of Multiple Deprivation [IMD]), socioeconomic factors (education level and employment status), health-related factors (presence of chronic conditions), lifestyle characteristics (smoking status and transport use), and uptake of other respiratory vaccinations (influenza and COVID-19) will additionally be defined using the profile survey. We did not investigate RSV vaccination given the available sample size, restricted eligibility for the study period and introduction of the programme during the study period.

Reasons provided for receiving or not receiving an influenza vaccination will additionally be derived from the profile questionnaire (see Table 1). These questions are asked to those who report having received or are planning to receive the vaccination that season and those who report no plan to receive the vaccine, respectively.

**Table 1.**
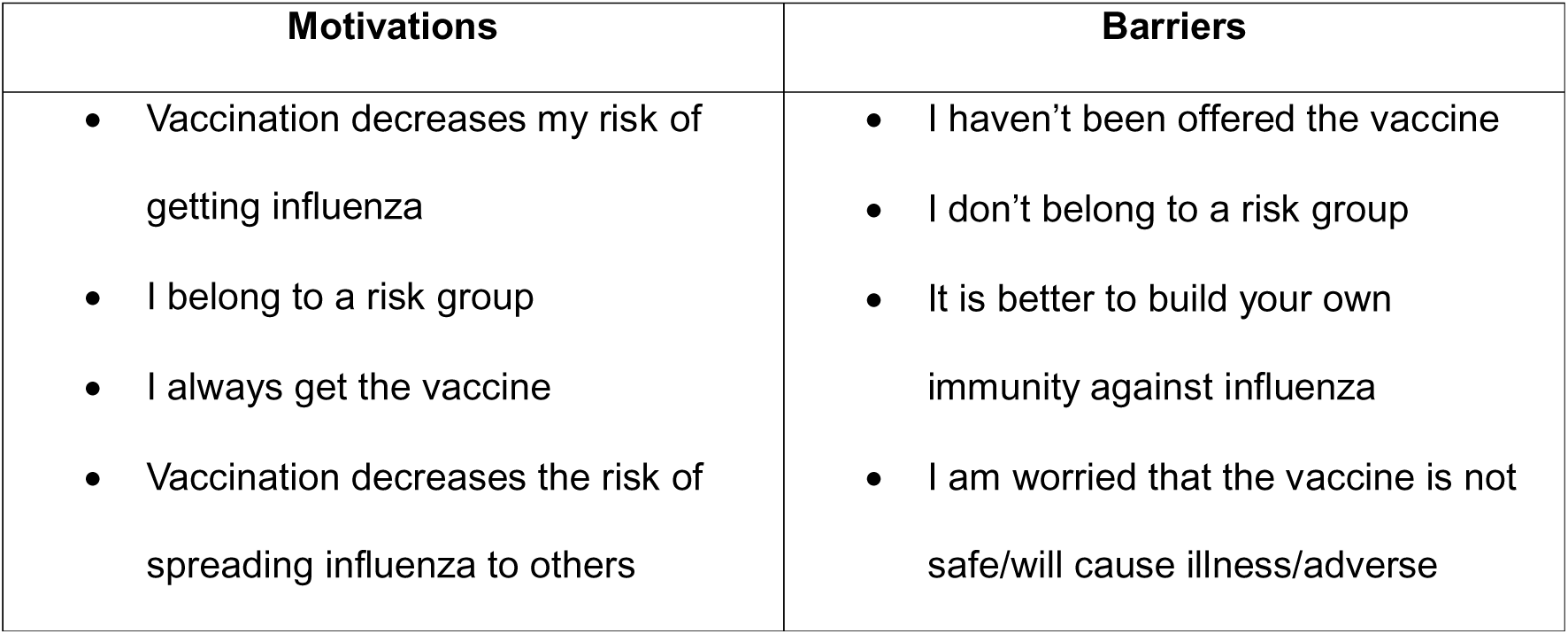
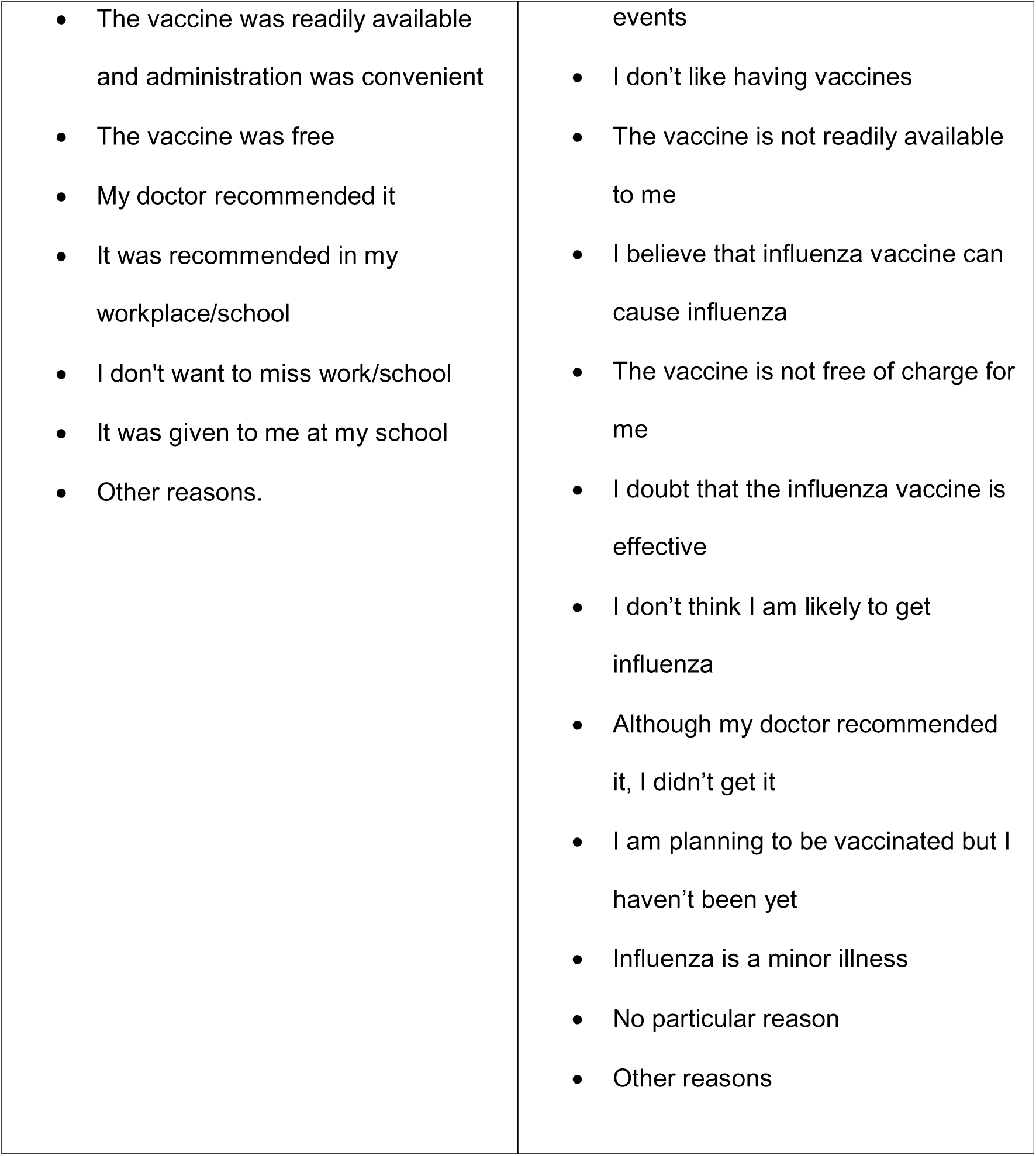
Reasons for receiving (or planning to receive) and not receiving the seasonal influenza vaccination included as response options in the FluSurvey background survey

Questions included in the profile survey and a table of how each factor was derived can be found in the supplementary material.

### 2.1 Statistical methods

All statistical analyses were conducted using R.

#### Participant Characteristics

Descriptive statistics were used to summarise the characteristics (country, sex, age group, education level, employment status, main mode of transport, IMD and ethnicity) of FluSurvey participants across the two seasons; where appropriate these proportions were compared to proportions from the 2021 census (12). Characteristics of FluSurvey participants vaccinated for either influenza or COVID-19 were also summarised.

#### Reasons for Receiving and Not Receiving an Influenza Vaccination

Exploratory descriptive analyses were conducted to examine self-reported reasons for receiving or not receiving an influenza vaccination across the two study seasons. Only those aged over 65 were included in this analysis to ensure eligibility wasn’t captured as a barrier. Proportions were calculated for each reason, and the most reported motivations and barriers were highlighted. Response options for motivations and barriers within in the questionnaire are presented in table 1.

#### Association Between Participant Characteristics and Vaccination Uptake

Associations between participant characteristics and vaccination uptake were assessed using logistic regression models. Analyses included participants aged 65 years and older and were conducted separately for influenza and COVID-19 vaccination uptake across the two winter seasons included in the study. Each vaccination outcome and season was modelled independently to allow for seasonal and vaccine-specific differences.

Vaccination status was treated as a binary outcome variable (vaccinated vs not vaccinated). Explanatory variables included sex, age group, receiving the other vaccination (COVID-19 or influenza), education level, employment status, main mode of transport, IMD, smoking, and number of chronic conditions. Due to small sample sizes in certain categories, levels of selected categorical variables were grouped to improve model convergence and precision of effect estimates (full details of factor grouping can be found in the supplementary material). The reference for each factor was chosen as the level with the largest sample size.

Initially, univariable logistic regression models were fitted for each explanatory variable to estimate crude associations with vaccination uptake. From each univariable model, crude odds ratios (ORs) with 95% confidence intervals (CIs) and corresponding p-values were derived.

Two multivariable logistic regression models were then constructed to assess the independent association between each factor and vaccination uptake while adjusting for potential confounding. Model one adjusts only for age and sex while model 2 adjusts for age, sex, IMD and education. Adjusted ORs with 95% CIs were reported in the main text.

## 3. Results

Table 1 presents the characteristics of FluSurvey participants across the 2023–24 (N=2,362) and 2024–25 (N=2,540) seasons, alongside 2021 UK census data for comparison (12). A table showing the characteristics of those over 65 is available in the supplementary material. Participants were predominantly resident in England (88% in both seasons), with smaller proportions from Scotland, Wales, and Northern Ireland.

Females were overrepresented in both seasons (63–64%) relative to the census population (51.1%). The age distribution was skewed toward middle-aged adults: individuals aged 45– 64 years comprised the largest group (40% in 2023–24; 42% in 2024–25), while those aged ≥85 years were underrepresented compared with census figures. Younger adults (≤44 years) were also somewhat underrepresented.

Participants were more highly educated than the general population, with 71–72% reporting a bachelor’s degree or higher compared with 33.6% in the census. In contrast, those with only GCSE/A-level qualifications or no formal education were underrepresented.

Regarding employment, a substantial proportion of respondents were in full- or part-time work (41–42%), slightly lower than census estimates (47.6%), while retired individuals were overrepresented (44–45% vs 21.6%). Other employment categories, including self-employment and unemployment, were broadly comparable or slightly lower than population estimates.

Car use was the dominant mode of transport (63% in both seasons), with smaller proportions reporting walking (17%), public transport use (15–16%), or cycling (≈5%). The sample was distributed across deprivation quintiles (IMD), with a modest skew toward less deprived groups (quintile 5: 29% in both seasons).

The cohort was predominantly of White ethnicity (97%), with minority ethnic groups underrepresented compared with census data. Overall, the FluSurvey sample differed from the general UK population, particularly in terms of sex, age distribution, educational attainment, and ethnicity, which should be considered when interpreting study findings.

A table describing the characteristics of FluSurvey participants who reported vaccination against influenza and COVID-19 in the 2023–24 and 2024–25 seasons can be found in the supplementary material. Overall, the distribution of vaccinated participants was broadly similar across both vaccines and seasons. In 2023-2024, 81% of participants over 65 reported receiving an Influenza vaccine and 73 % reported receiving a COVID vaccine. In 2024-2025, 81% of participants over 65 reported receiving an Influenza vaccine and 83% reported receiving a COVID vaccine.

Figure 1 shows the reasons given for getting vaccinated, among those aged 65 and above. The most popular reason given in each season was “I belong to a risk group” followed by “decreases the risk of getting influenza”. The least popular reasons given were those related to work and school. This is to be expected in this sample as they are all over 65 and the majority are retired.

**Figure 1.**
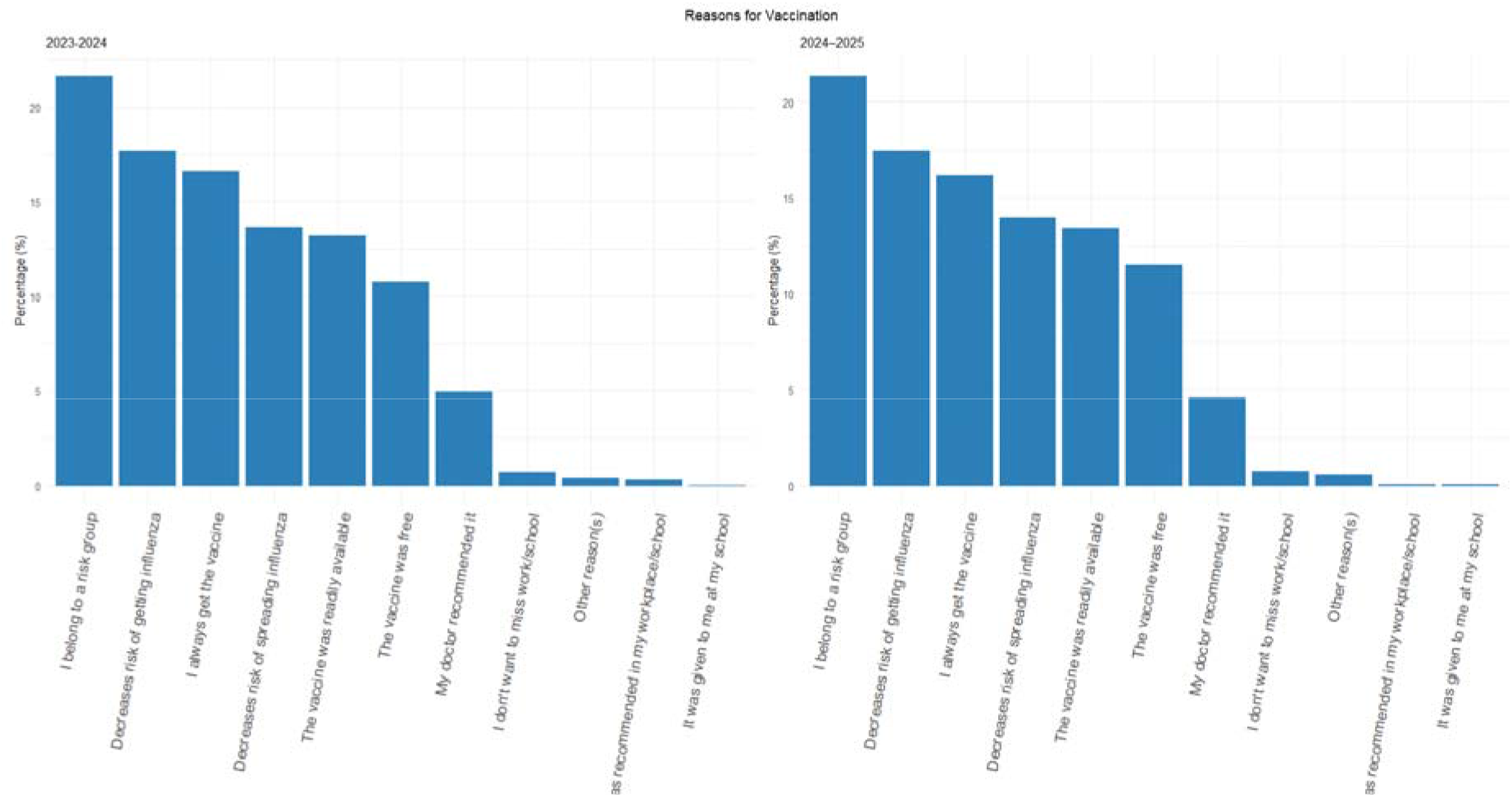
Reasons for influenza vaccination reported by participants during the 2023–2024 and 2024–2025 seasons. The bar charts display the percentage of respondents endorsing each reason, highlighting the most and least common motivations.

Figure 2 shows the reasons people gave for not getting vaccinated. The three most popular reasons, “It is better to build natural immunity to influenza”, “I am worried that the vaccine is not safe/will cause illness/other adverse effects” and “other reasons” were the same across the two seasons with some fluctuation in positioning. In 2023-2024 season the least common reason for not being vaccinated was “I haven’t been offered the vaccine”, but “the vaccine is not readily available to me” and “I’m planning to be vaccinated but haven’t been yet” were never selected as reasons. In 2024 the least popular reason was “I don’t think I’m likely to get influenza”. Again, several options were never selected as reasons; “influenza is a minor illness”, “the vaccine is not readily available to me”, “The vaccine is not free” and “I’m planning to be vaccinated but haven’t been yet”.

**Figure 2.**
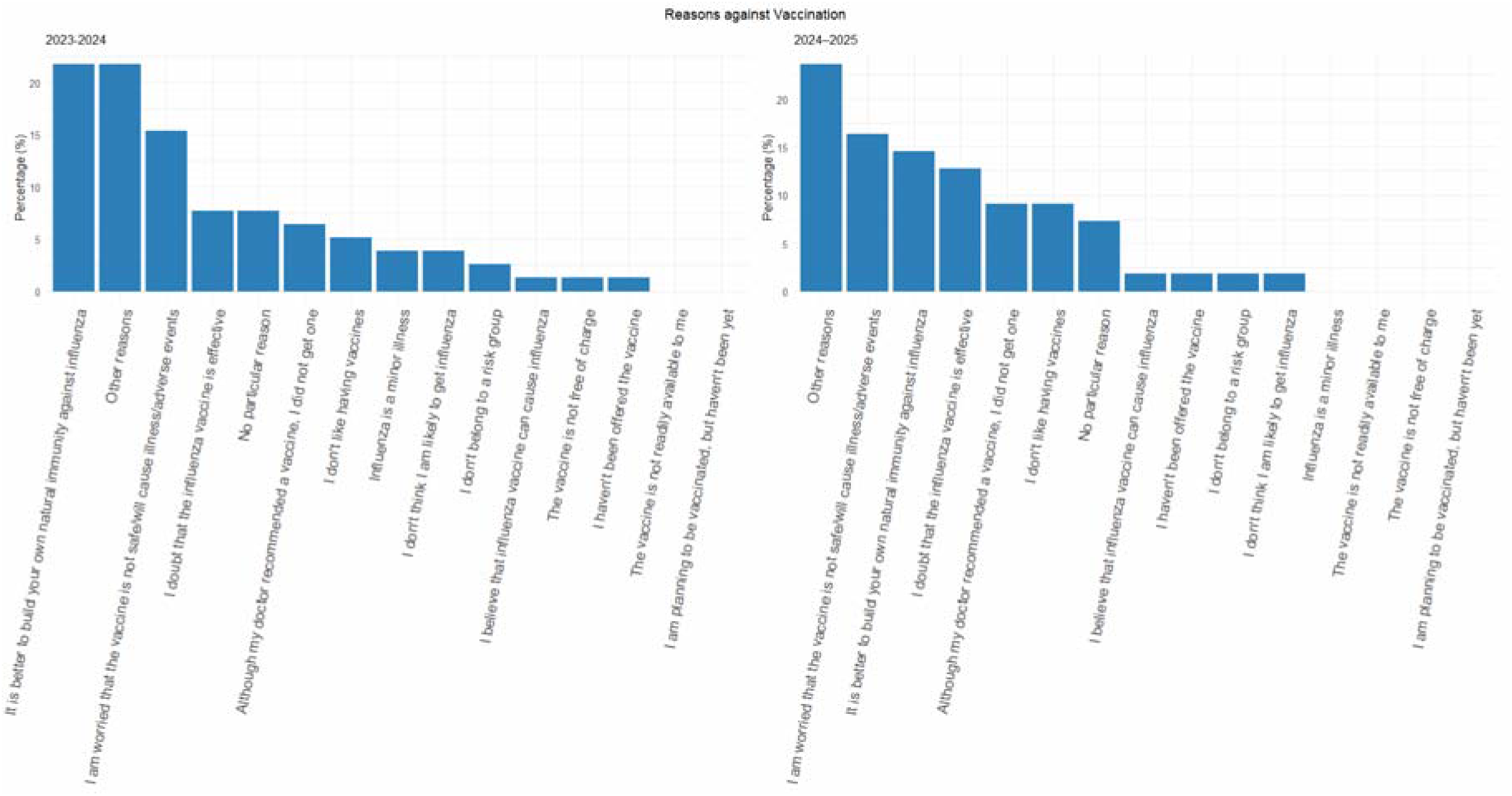
Reasons against influenza vaccination reported by participants during the 2023–2024 and 2024–2025 seasons. The bar charts display the percentage of respondents endorsing each reason, highlighting the most and least common motivations.

### Factors associated with influenza vaccination

Tables 3 and 4 present logistic regression analyses of factors associated with influenza vaccination uptake among adults aged ≥65 years across the 2023–2024 and 2024–2025 seasons. Table 3 shows results adjusted for age and sex while Table 4 shows results adjusted for age, sex, IMD and education. Results of an unadjusted regression are presented in the supplementary material.

**Table 2.** Characteristics of FluSurvey participants from 2023–24 to 2024–25 season with national census 2021 UK data for comparison.

|  | 2023-24<br>(N=2,362) <sup>1</sup> | 2024-25<br>(N=2,540) <sup>1</sup> | Census 2021 (%)<sup>2</sup> |
| --- | --- | --- | --- |
| Country |  |  |  |
| England | 2,237 (88%) | 2,072 (88%) | 84.4% |

|  |  |  | Census 2021 (%)<sup>2</sup> |
| --- | --- | --- | --- |
|  | 2023-24<br>(N=2,362)<sup>1</sup> | 2024-25<br>(N=2,540)<sup>1</sup> |  |
| Scotland | 165 (6.5%) | 143 (6.1%) | 8.1% |
| Wales | 81 (3.2%) | 88 (3.7%) | 4.6% |
| Northern Ireland | 14 (0.6%) | 11 (0.5%) | 2.8% |
| Other | 6 (0.2%) | 6 (0.2%) |  |
| **Sex** |  |  |  |
| Female | 1,595 (63%) | 1,523 (64%) | 51.1% |
| Male | 932 (37%) | 831 (35%) | 48.9% |
| Other | 13 (0.5%) | 8 (0.3%) |  |
| **Age group** |  |  |  |
| <18 | 51 (2.0%) | 34 (1.4%) | 21.8% |
| 19-44 | 298 (12%) | 315 (13%) | 33.6% |
| 45-64 | 1,021 (40%) | 981 (42%) | 12.8% |
| 65-74 | 766 (30%) | 674 (29%) | 10.01% |
| 75-84 | 356 (14%) | 327 (14%) | 6.2% |
| >85 | 41 (1.6%) | 29 (1.2%) | 2.4% |
| Unknown | 7 | 2 |  |
| **Education** |  |  |  |
| Bachelors degree or higher | 1,799 (71%) | 1,690 (72%) | 33.6% |
| GCSE / A-level or equivalent | 633 (25%) | 596 (25%) | 48.1% |
| No formal education | 69 (2.7%) | 51 (2.2%) | 18.3% |
| Still in education | 39 (1.5%) | 25 (1.1%) |  |
| **Employment status** |  |  |  |
| Daycare or education | 72 (2.8%) | 43 (1.8%) |  |
| Full or part-time employment | 1,044 (41%) | 1,000 (42%) | 47.6% |
| Homemaker | 61 (2.4%) | 69 (2.9%) | 4.8% |
| Long-term sick or parental leave | 45 (1.8%) | 55 (2.3%) | 4.2% |
| Other | 34 (1.3%) | 34 (1.4%) | 3.1% |
| Retired | 1,138 (45%) | 1,034 (44%) | 21.6% |
| Self-employment | 128 (5.0%) | 112 (4.7%) | 9.6% |
| Unemployed | 18 (0.7%) | 15 (0.6%) | 3.4% |
| **Transport** |  |  |  |
| Bike | 122 (4.8%) | 109 (4.6%) |  |
| Car | 1,593 (63%) | 1,488 (63%) |  |
| Other | 21 (0.8%) | 25 (1.2%) |  |
|  | 2023-24<br>(N=2,362)<sup>1</sup> | 2024-25<br>(N=2,540)<sup>1</sup> |  |
| Public transport | 376 (15%) | 349 (15%) |  |
| Walking | 428 (17%) | 391 (17%) |  |
| **IMD** |  |  |  |
| 1 | 217 (11%) | 256 (11%) |  |
| 2 | 339 (17%) | 416 (18%) |  |
| 3 | 396 (20%) | 438 (19%) |  |
| 4 | 441 (22%) | 550 (24%) |  |
| 5 | 576 (29%) | 663 (29%) |  |
| Unknown | 571 | 39 |  |
| **Ethnicity** |  |  |  |
| Other ethnic groups<sup>3</sup> |  | 71 (3.2%) | 16.3% |
| White |  | 2,205 (97%) | 83.1% |
| Unknown |  | 86 |  |

<sup>1</sup>Values are n (%). Percentages may not sum to 100 due to rounding.
<sup>2</sup>Census 2021 England and Wales data used for employment status as UK data was not available.
Questions about participant ethnicity were added to FluSurvey in 2024-25 so ethnicity data is not available for the 2023-24 season.
<sup>3</sup> The following ethnic groups have been grouped due to small sample sizes Asian, Asian British or Asian Welsh, Black, Black British, Black Welsh, Caribbean or Africa, Mixed or Multiple ethnic groups

**Table 3.** Factors associated with influenza vaccination uptake (age 65+; n = 961 in 2023-2024 and n = 840 in 2024-2025). Odds ratios with 95% confidence intervals adjusted for age and sex are presented. Statistically significant associations (p<0.05) are highlighted in blue.

| Predictor | Level | 2023-2024 |  |  | 2024-2025 |  |  |
| --- | --- | --- | --- | --- | --- | --- | --- |
|  |  | OR | 95% CI | p-value | OR | 95% CI | p-value |
| Sex | Female (Ref) | 1 | - | - | 1 | - | - |
|  | Male | 1.03 | 0.76–1.42 | 0.832 | 0.86 | 0.63–1.2 | 0.375 |
| Age group | 65-74 (Ref) | 1 | - | - | 1 | - | - |
|  | 75-84 | 1.02 | 0.73–1.44 | 0.895 | 0.67 | 0.48–0.94 | 0.020 |
|  | >85 | 0.64 | 0.32–1.41 | 0.239 | 0.72 | 0.3–1.99 | 0.488 |
| IMD quintile | 5 (Ref) | 1 | - | - | 1 | - | - |
|  | 1-2 | 1.33 | 0.84–2.13 | 0.225 | 0.89 | 0.59–1.36 | 0.601 |
|  | 3-4 | 1.25 | 0.84–1.86 | 0.266 | 1.18 | 0.81–1.72 | 0.381 |
| Education | Degree or above (Ref) | 1 | - | - | 1 | - | - |
|  | High school or below | 0.72 | 0.52–0.99 | 0.040 | 0.76 | 0.54–1.07 | 0.111 |
| Employment status | Not employed (Ref) | 1 | - | - | 1 | - | - |
|  | Employed | 1.25 | 0.79–2.06 | 0.361 | 1.59 | 0.93–2.88 | 0.106 |
| Transport | Not public transport (Ref) | 1 | - | - | 1 | - | - |
|  | Public transport | 1.19 | 0.73–2.06 | 0.500 | 0.52 | 0.34–0.82 | 0.004 |
| COVID-19 vaccination | No (Ref) | 1 | - | - | 1 | - | - |
|  | Yes | 16.66 | 11.58–24.35 | <0.001 | 10.16 | 6.95–14.96 | <0.001 |
| Smoking | No (Ref) | 1 | - | - | 1 | - | - |
|  | Yes | 0.52 | 0.26–1.11 | 0.074 | 0.46 | 0.24–0.94 | 0.025 |
| Chronic Condition | None (Ref) | 1 | - | - | 1 | - | - |
|  | More than 1 | 1.00 | 0.56–1.9 | 0.996 | 0.88 | 0.49–1.67 | 0.688 |
|  | One | 0.77 | 0.55–1.09 | 0.137 | 0.96 | 0.67–1.4 | 0.824 |

**Table 4.** Factors associated with influenza vaccination uptake (age 65+; n = 961 in 2023-2024 and n = 840 in 2024-2025). Logistic regression odds ratios with 95% confidence intervals adjusted for age, sex IMD and education. Statistically significant associations (p<0.05) are highlighted in blue.

| Predictor | Level | 2023-2024 |  |  | 2024-2025 |  |  |
| --- | --- | --- | --- | --- | --- | --- | --- |
|  |  | OR | 95% CI | p-value | OR | 95% CI | p-value |
| Employment status | Not employed (Ref) | 1 | - | - | 1 | - | - |
|  | Employed | 1.46 | 0.81–2.76 | 0.228 | 1.78 | 1–3.37 | 0.062 |
| Transport | Not public transport (Ref) | 1 | - | - | 1 | - | - |
|  | Public transport | 0.86 | 0.46–1.7 | 0.661 | 0.46 | 0.29–0.76 | 0.002 |
| COVID-19 vaccination | No (Ref) | 1 | - | - | 1 | - | - |
|  | Yes | 13.90 | 9.28–21.17 | <0.001 | 8.54 | 5.82–12.6 | <0.001 |
| Smoking | No (Ref) | 1 | - | - | 1 | - | - |
|  | Yes | 0.33 | 0.12–0.94 | 0.033 | 0.51 | 0.24–1.12 | 0.081 |
| Chronic Condition | None (Ref) | 1 | - | - | 1 | - | - |
|  | More than 1 | 0.74 | 0.35–1.66 | 0.445 | 0.91 | 0.47–1.84 | 0.776 |
|  | One | 0.75 | 0.48–1.18 | 0.212 | 0.91 | 0.6–1.38 | 0.640 |
| IMD quintile | 5 (Ref) | 1 | - | - | 1 | - | - |
|  | 1-2 | 1.37 | 0.81–2.35 | 0.249 | 1.08 | 0.68–1.74 | 0.746 |
|  | 3-4 | 1.12 | 0.71–1.77 | 0.632 | 1.28 | 0.85–1.94 | 0.240 |
| Education | Degree or above (Ref) | 1 | - | - | 1 | - | - |
|  | High school or below | 0.94 | 0.62–1.45 | 0.789 | 0.87 | 0.6–1.27 | 0.461 |

Prior COVID-19 vaccination was strongly associated with influenza vaccine uptake in both seasons and models, with substantially increased odds of vaccination (model 1 OR=16.66 [11.58-24.35] in 2023-2024; OR=10.16 [6.95-14.96] in 2024-2025), (model 2 OR=13.90 [9.28-21.17] in 2023-2024; OR=8.54 [5.82-12.60] in 2024-2025). Sex, IMD, employment status and chronic conditions were not strongly associated with vaccine uptake in either season or model, although we note we lacked statistical power to detect smaller effects.

Some variation was observed across seasons and by model. Individuals aged 75–84 years had lower odds of vaccination in 2024–2025 (OR=0.67 [0.48-0.94]). Educational attainment showed a modest association in 2023–2024 model 1, with individuals educated to high school level or below having lower odds of influenza vaccination compared with high school level and above (OR=0.72 [0.52-0.99]). However, this relationship attenuated following adjustment for IMD. Public transport use was associated with significantly lower odds of vaccination in 2024–2025 only (model 1 OR=0.52 [0.34-0.82], model 2 OR=0.46 [0.29- 0.76]). Smoking status was associated with reduced uptake in both seasons, although confidence intervals included the null in some models), (model 1 OR=0.52 [0.26-1.11], model 2 OR=0.33 [0.12-0.94] in 2023-2024), (model 1 OR=0.46 [0.24-0.94], model 2 OR=0.51 [0.24-1.12] in 2024-2025).

Figure 3 shows a visual comparison of the two models across the two study seasons for influenza vaccination uptake.

**Figure 3.**
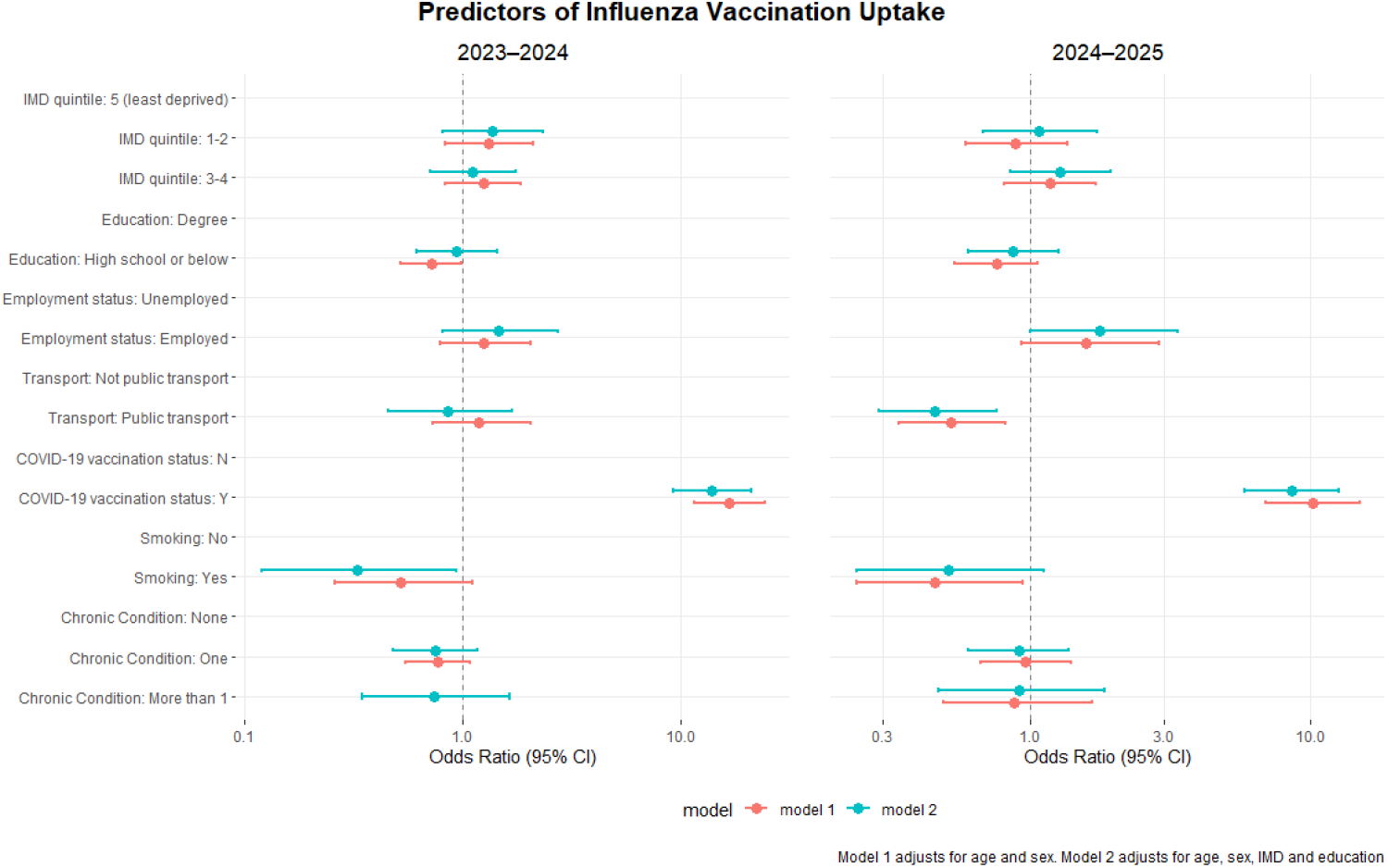
A forest plot presenting results from analyses investigating factors associated with influenza vaccine uptake. Model 1 (orange) adjusts for age and sex whereas model 2 (blue) adjusts for age, sex, IMD and education. Reference groups for each predictor are included below for context, with no estimate plotted.

#### Factors associated with COVID-19 Vaccination

Tables 5 and 6 present logistic regression analyses of factors associated with COVID-19 vaccination uptake among adults aged ≥65 years across the 2023–2024 and 2024–2025 seasons. Table 5 shows results adjusted for age and sex while Table 6 shows results adjusted for age, sex, IMD and education. Results of an unadjusted regression are presented in the supplementary material.

**Table 5.**
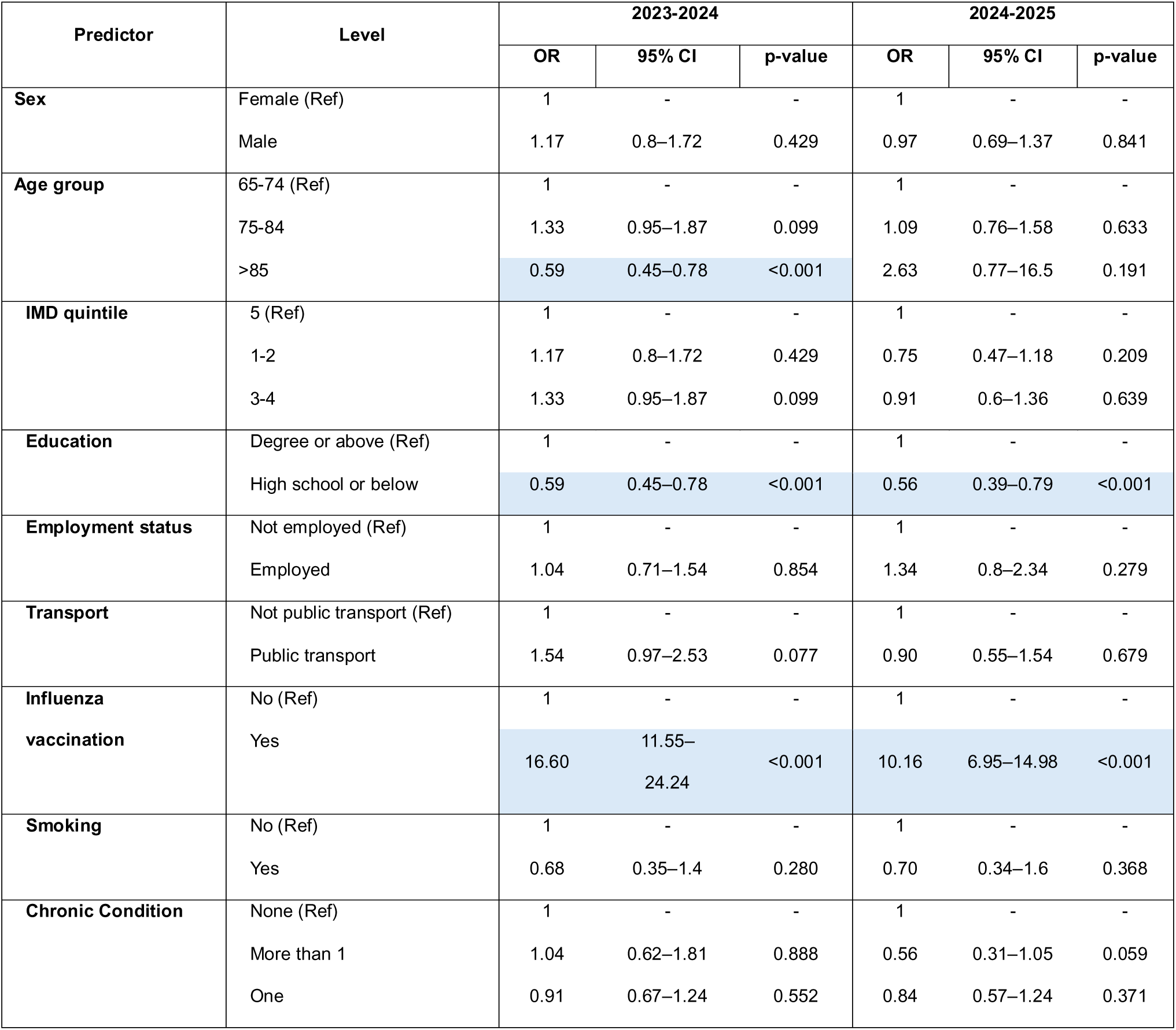
Factors associated with COVID-19 vaccination uptake (age 65+; n = 857 in 2023-2024 and n = 859 in 2024-2025). Odds ratios with 95% confidence intervals adjusted for age and sex are presented. Statistically significant associations (p<0.05) are highlighted in blue.

**Table 6.**
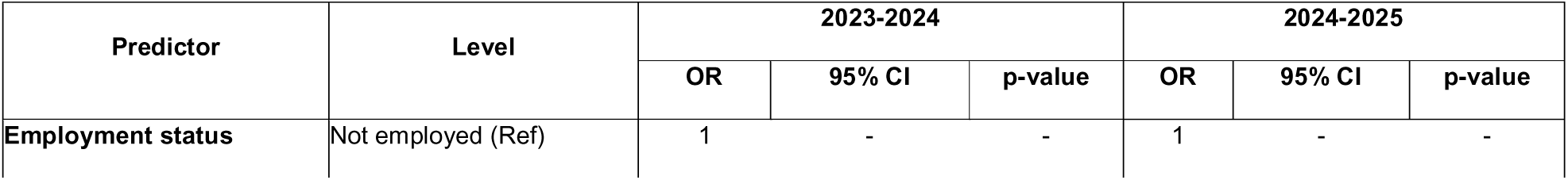
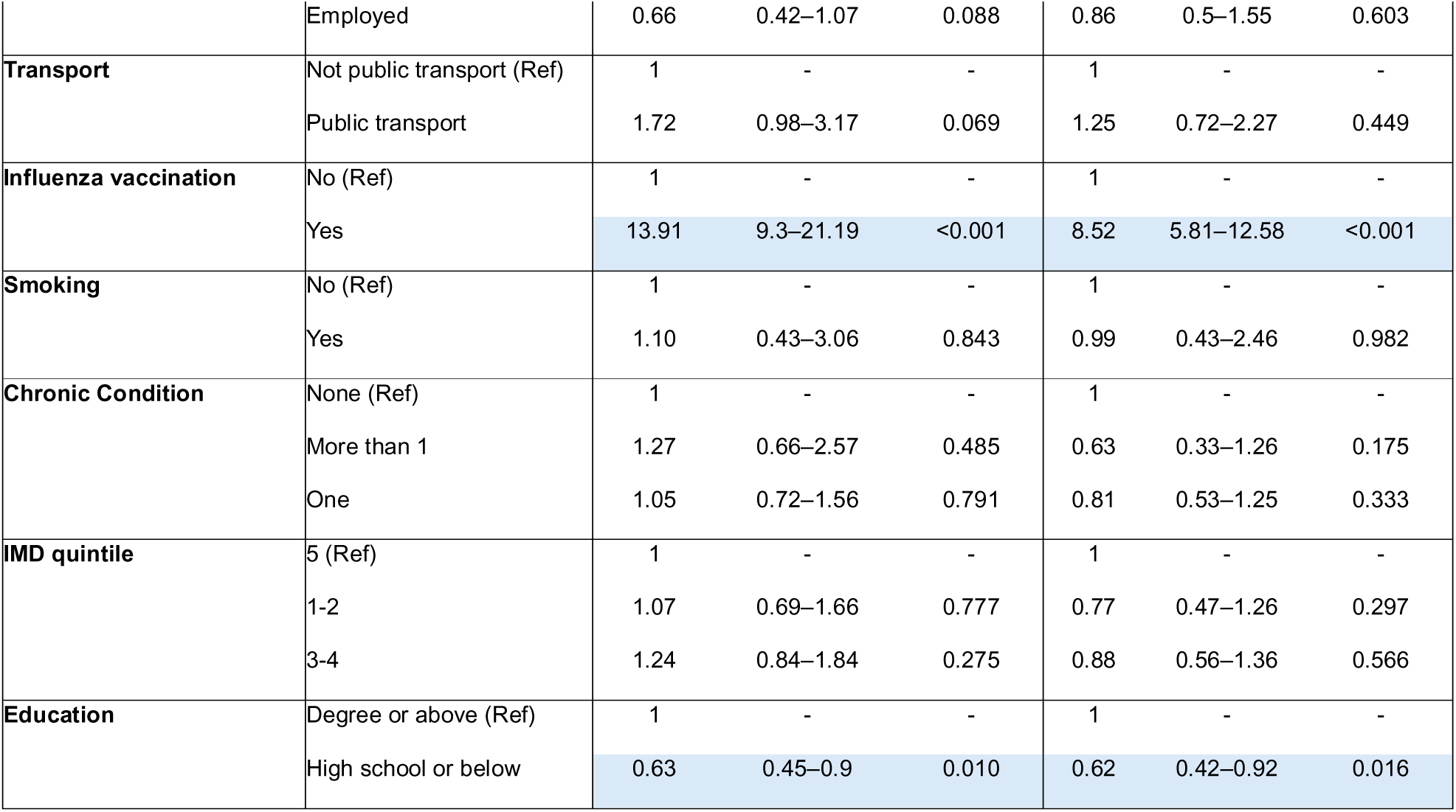
Factors associated with COVID-19 vaccination uptake (age 65+; n = 857 in 2023-2024 and n = 859 in 2024-2025). Odds ratios with 95% confidence intervals adjusted for age and sex are presented. Statistically significant associations (p<0.05) are highlighted in blue.

| Predictor | Level | 2023-2024 |  |  | 2024-2025 |  |  |
| --- | --- | --- | --- | --- | --- | --- | --- |
|  |  | OR | 95% CI | p-value | OR | 95% CI | p-value |
| Employment status | Not employed (Ref) | 1 | - | - | 1 | - | - |
|  | Employed | 0.66 | 0.42–1.07 | 0.088 | 0.86 | 0.5–1.55 | 0.603 |
| **Transport** | Not public transport (Ref) | 1 | - | - | 1 | - | - |
|  | Public transport | 1.72 | 0.98–3.17 | 0.069 | 1.25 | 0.72–2.27 | 0.449 |
| **Influenza vaccination** | No (Ref) | 1 | - | - | 1 | - | - |
|  | Yes | 13.91 | 9.3–21.19 | <0.001 | 8.52 | 5.81–12.58 | <0.001 |
| **Smoking** | No (Ref) | 1 | - | - | 1 | - | - |
|  | Yes | 1.10 | 0.43–3.06 | 0.843 | 0.99 | 0.43–2.46 | 0.982 |
| **Chronic Condition** | None (Ref) | 1 | - | - | 1 | - | - |
|  | More than 1 | 1.27 | 0.66–2.57 | 0.485 | 0.63 | 0.33–1.26 | 0.175 |
|  | One | 1.05 | 0.72–1.56 | 0.791 | 0.81 | 0.53–1.25 | 0.333 |
| **IMD quintile** | 5 (Ref) | 1 | - | - | 1 | - | - |
|  | 1-2 | 1.07 | 0.69–1.66 | 0.777 | 0.77 | 0.47–1.26 | 0.297 |
|  | 3-4 | 1.24 | 0.84–1.84 | 0.275 | 0.88 | 0.56–1.36 | 0.566 |
| **Education** | Degree or above (Ref) | 1 | - | - | 1 | - | - |
|  | High school or below | 0.63 | 0.45–0.9 | 0.010 | 0.62 | 0.42–0.92 | 0.016 |

Prior influenza vaccination was strongly associated with COVID-19 vaccine uptake in both seasons and models, with substantially increased odds of vaccination (model 1 OR=16.60 [11.55-24.24] in 2023-2024; OR=10.16 [6.95-14.98] in 2024-2025), (model 2 OR=13.91 [9.30-21.19] in 2023-2024; OR=8.52 [5.81-12.58] in 2024-2025). Lower educational attainment was associated with decreased odds of COVID-19 vaccination in both seasons and models (model 1 OR=0.59 [0.45-0.78] in 2023-2024; OR=0.56 [0.39-0.79] in 2024- 2025), (model 2 OR=0.63 [0.45-0.90] in 2023-2024; OR=0.62 [0.42-0.92] in 2024-2025).

Sex, IMD, employment status, chronic conditions and transport were not strongly associated with vaccine uptake in either season or model, although we note we lacked statistical power to detect smaller effects.

Being >85 years was associated with decreased odds of vaccination in model 1 in 2023- 2024 (OR=0.59 [0.45-0.78]). However, there were few in this age group and there was no evidence of an association in any other season or model. Figure 4 shows a visual comparison of the two models across the two study seasons for COVID-19 vaccination uptake.

**Figure 4.**
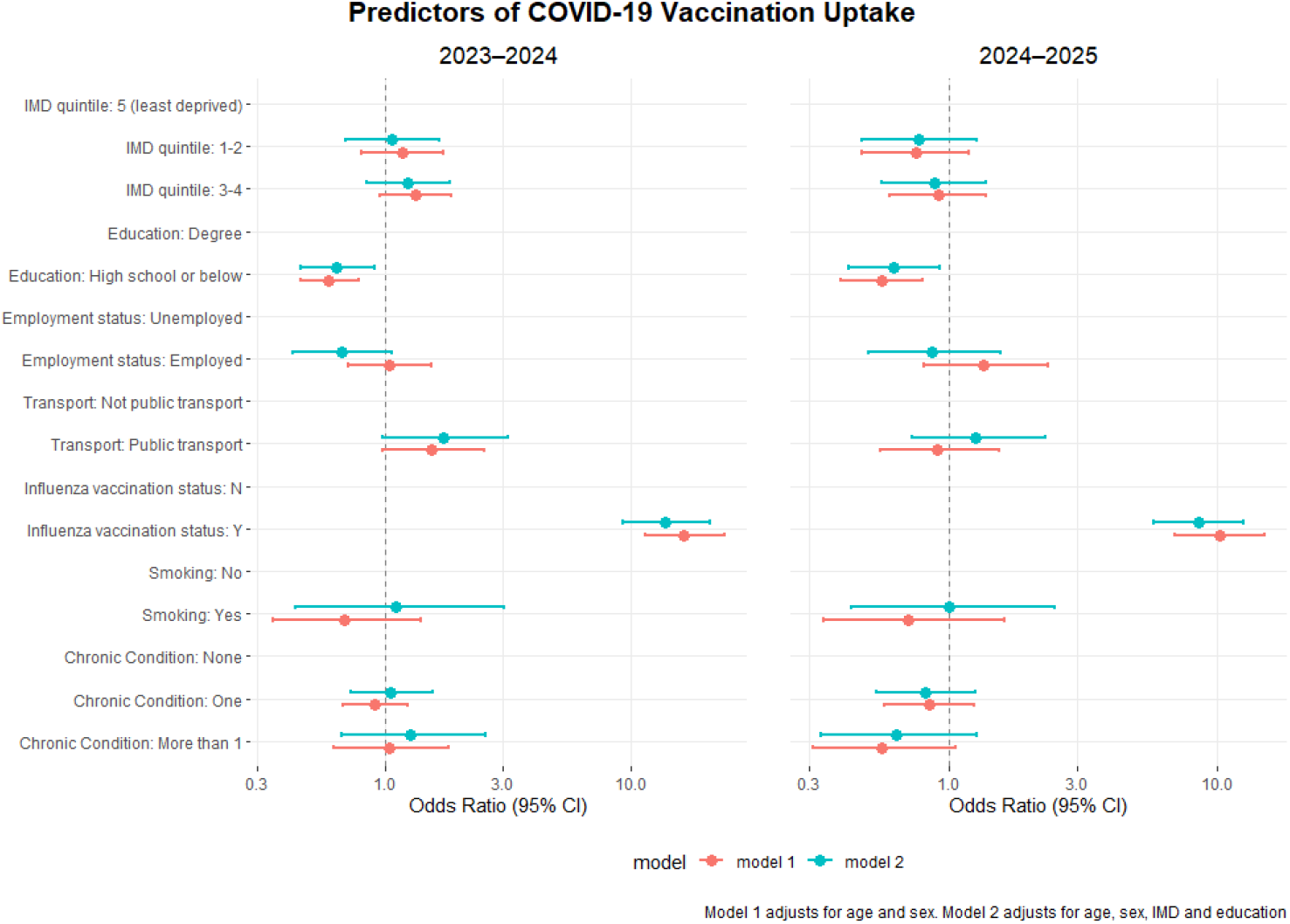
A forest plot presenting results from analyses investigating factors associated with COVID-19 vaccine uptake. Model 1 (orange) adjusts for age and sex whereas model 2 (blue) adjusts for age, sex, IMD and education. Reference groups for each predictor are included below for context, with no estimate plotted.

## 4. Discussion

In this study of adults aged ≥65 years participating in FluSurvey, self-reported vaccination uptake was high across both seasons, with 81% reporting influenza vaccination in both 2023–2024 and 2024–2025, and COVID-19 vaccination uptake increasing from 73% to 83%. These estimates exceed national routine data for COVID-19 vaccination and are broadly comparable, or slightly higher, for influenza vaccination in this age group. This likely reflects selection into FluSurvey, with overrepresentation of individuals who are older, more health conscious, and more highly educated, all of which are associated with higher vaccination uptake. Similar patterns of elevated uptake have been observed in other participatory surveillance systems, suggesting that these cohorts may better reflect individuals who are already engaged with their health and preventative behaviours.

In terms of behavioural drivers, perceived personal risk (“I belong to a risk group”) and perceived effectiveness (“decreases my risk of getting influenza”) were the dominant motivations for influenza vaccination, while concerns around safety and beliefs in natural immunity were the most frequently cited barriers. These findings align with previous literature on vaccine hesitancy, including recent COVID-19 studies, where safety concerns and perceived necessity consistently emerge as key determinants of uptake. In contrast, structural barriers such as access or cost were rarely reported, which may reflect the accessibility of vaccination services for this age group in the UK.

A key finding was the strong and consistent association between influenza and COVID-19 vaccination uptake. Individuals who received one vaccine were substantially more likely to receive the other across both seasons and models. This supports the concept that vaccine acceptance is not vaccine-specific but reflects a broader behavioural tendency. This observation is consistent with recent large-scale cohort evidence (9), which has shown that prior vaccination behaviour and general vaccine attitudes are among the strongest predictors of subsequent uptake. In the English context, where co-administration of influenza and COVID-19 vaccines is recommended among eligible groups, this finding has practical implications, suggesting that integrated delivery strategies may help maximise uptake across both programmes.

We observed limited and inconsistent associations between most sociodemographic characteristics and vaccination uptake. In contrast to some previous studies, we did not identify strong associations with deprivation (IMD), which is a well-established predictor of vaccine uptake. This may reflect limited statistical power, the need to group IMD categories, and reduced variability within this sample. Similarly, we did not observe an association between smoking and COVID-19 vaccination, nor consistent associations with comorbidity burden, both of which have been reported elsewhere (9). However, given the relatively small sample size and wide confidence intervals, these null findings should be interpreted cautiously.

Educational attainment was the most consistent sociodemographic factor associated with vaccination behaviour. Lower education was associated with reduced COVID-19 vaccination uptake across both seasons and models, in line with previous research (9). A similar but weaker association was observed for influenza vaccination, which attenuated after adjustment for IMD, suggesting potential confounding by deprivation. These findings highlight the need for further investigation in larger and more representative cohorts, particularly to better understand the interplay between socioeconomic factors and vaccination behaviour.

Some associations varied by season, including the observed relationship between public transport use and lower influenza vaccination uptake in 2024–2025, and a potential association with smoking. These findings are exploratory and may reflect residual confounding, behavioural differences, or chance variation. However, they are consistent with the broader literature suggesting that vaccination behaviours are context-dependent and may shift over time in response to changing public health messaging, perceived risk, and healthcare access. The longitudinal design of FluSurvey is therefore a key strength, offering the potential to monitor how these behaviours evolve across seasons.

This study has several strengths and limitations. It provides timely insights into vaccination attitudes and behaviours in the post-COVID-19 context, including both influenza and COVID- 19 vaccines within the same cohort. The availability of self-reported reasons for vaccination decisions adds important behavioural context that is often missing from routine data sources. However, the study is limited by a relatively small and non-representative sample, with overrepresentation of older, White, and highly educated individuals. This limits generalisability and may attenuate observed associations with key sociodemographic factors such as deprivation and ethnicity. Additionally, the need to group variables and the absence of adjustment for multiple comparisons may have reduced precision and increased the risk of both type I and type II error. Finally, attitudes toward COVID-19 vaccination were not captured, limiting direct comparison of behavioural drivers across vaccines.

Overall, these findings reinforce the importance of general vaccine acceptance as a key predictor of uptake, while suggesting that some traditional sociodemographic inequalities may be less apparent in this cohort. They also highlight the continued importance of addressing risk perception and safety concerns in vaccination campaigns. Future work using larger and more representative samples, and leveraging the longitudinal nature of FluSurvey, will be important to better understand how vaccination behaviours change over time and to inform targeted public health interventions.

## Data Availability

Participant data are unable to be shared; participants accept a privacy notice on registration which does not permit the sharing of pseudonymized data for this purpose (<u>UKHSAFluSurvey Privacy Notice | Flusurvey</u>, accessed 25/08/2026). For specific data requests, please refer to https://www.gov.uk/government/publications/accessing-ukhsa-protected-data.

## Data Availability

Participant data are unable to be shared; participants accept a privacy notice on registration which does not permit the sharing of pseudonymized data for this purpose (UKHSA FluSurvey Privacy Notice | Flusurvey, accessed 25/08/2026). For specific data requests, please refer to https://www.gov.uk/government/publications/accessing-ukhsa-protected-data.

## Supplementary Material

### S1. Survey phrasing and factor grouping

**Table S1.**
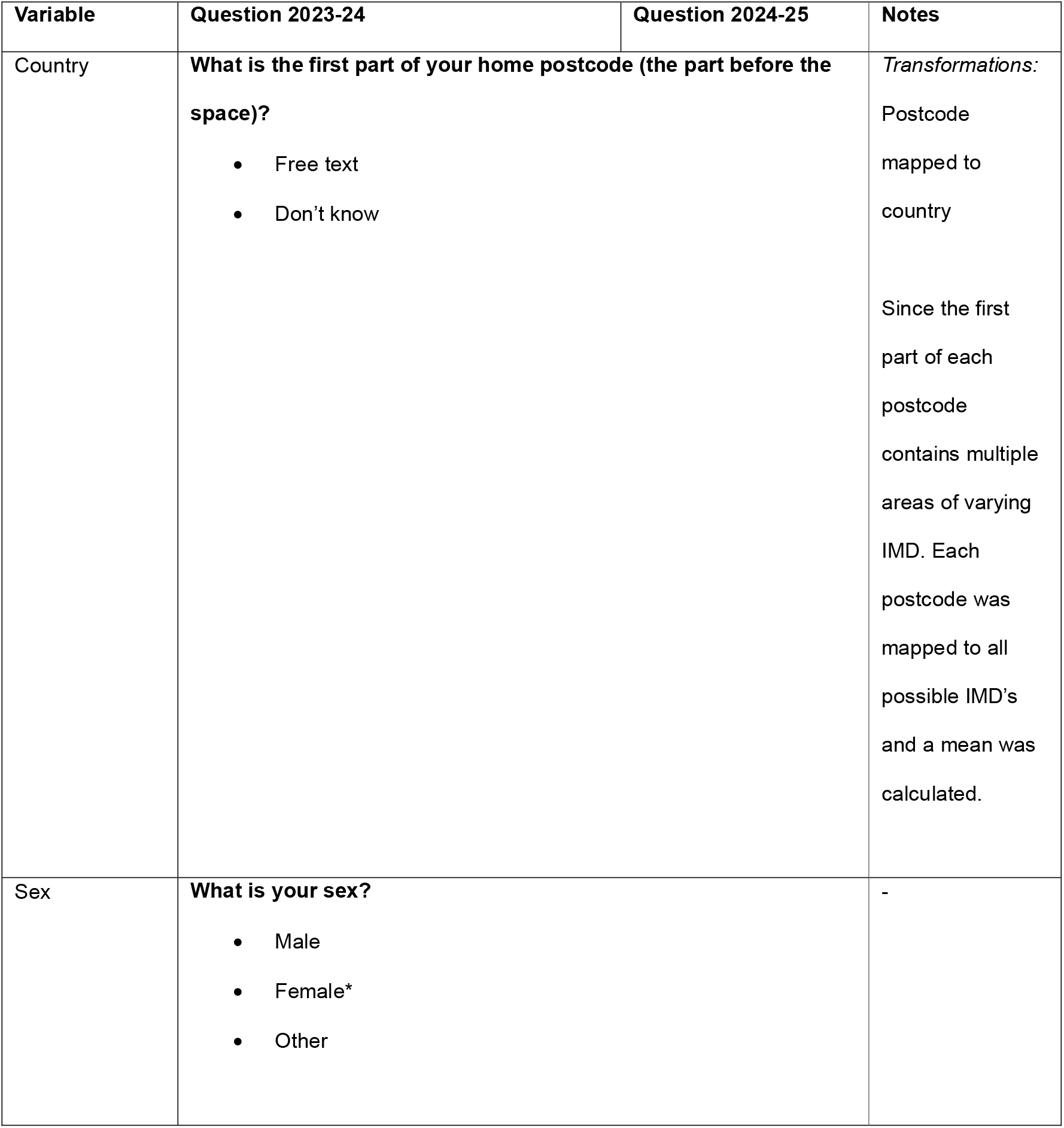
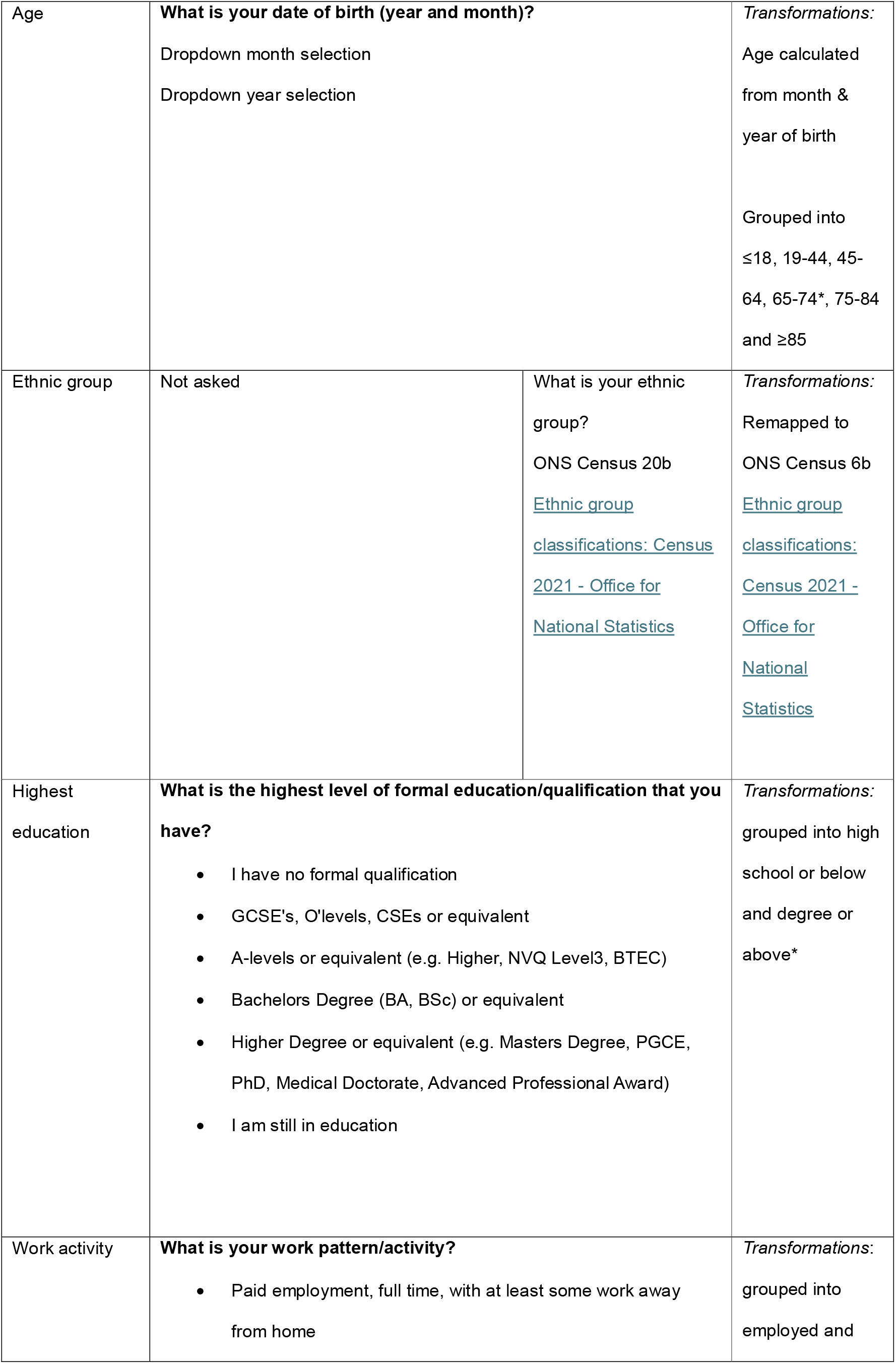
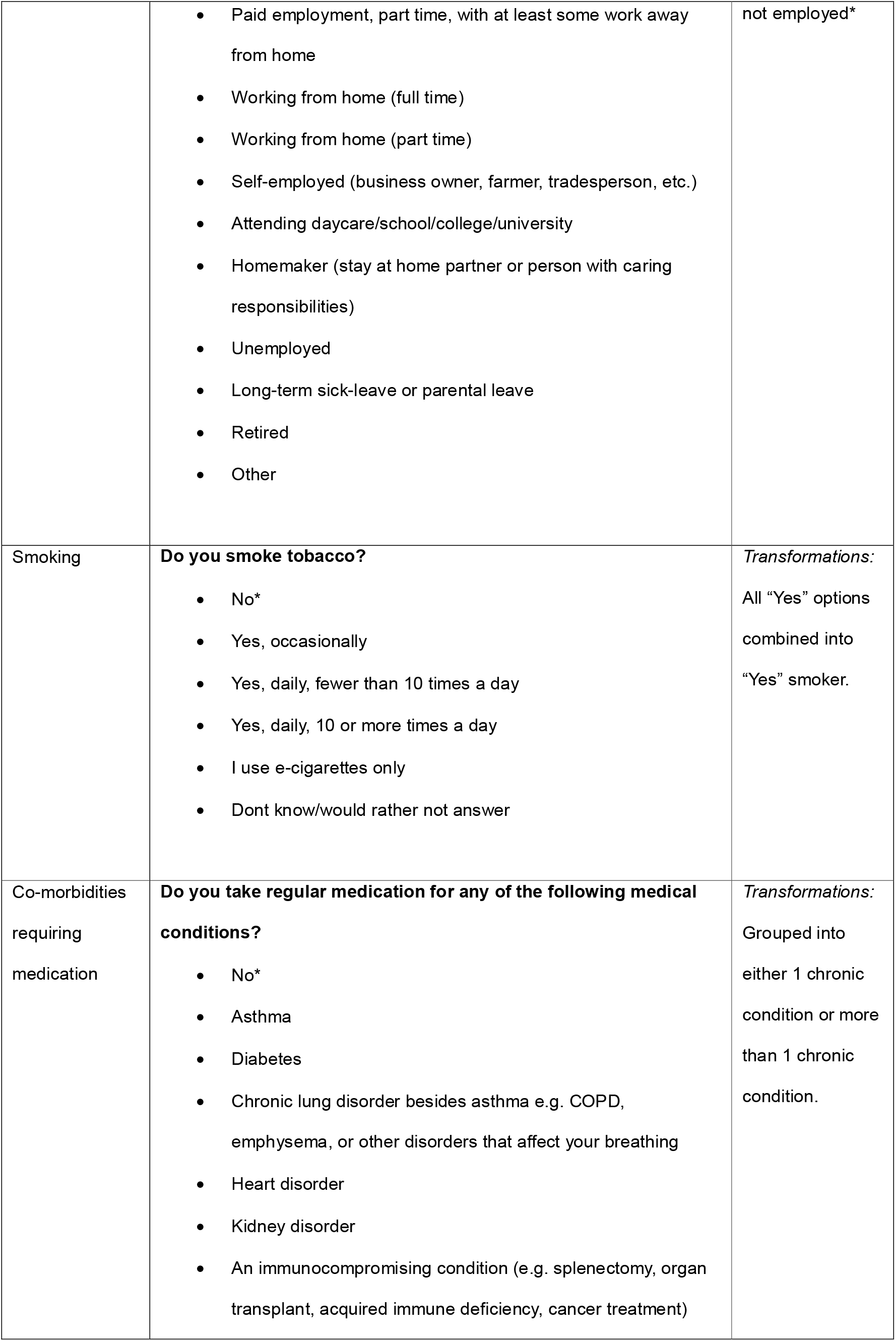
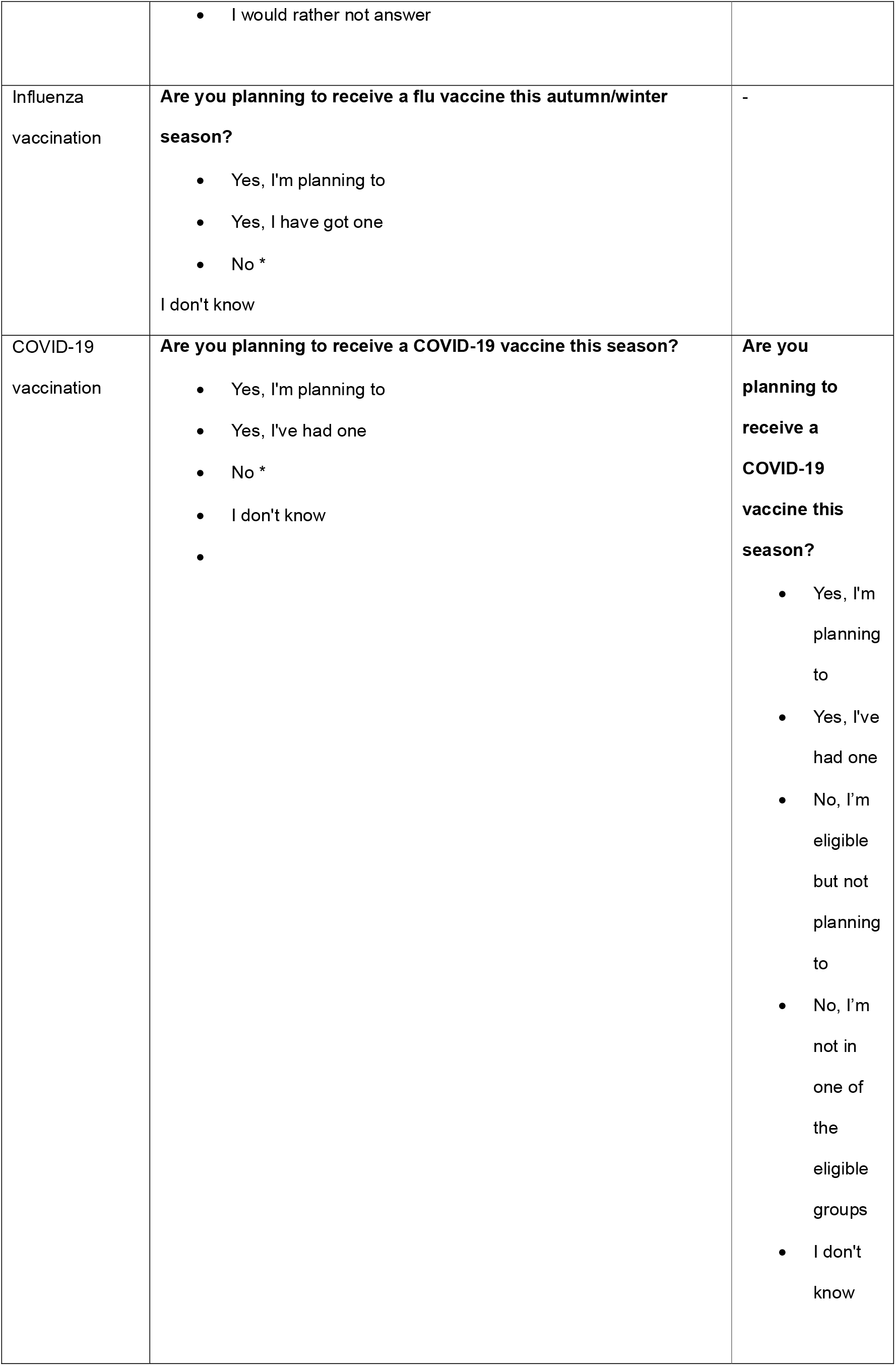
Survey phrasing for the variables derived in the present analyses are included below. Reference levels are indicated by an Asterix *.

### S2. Characteristics of participants who are over 65

**Table S2.**
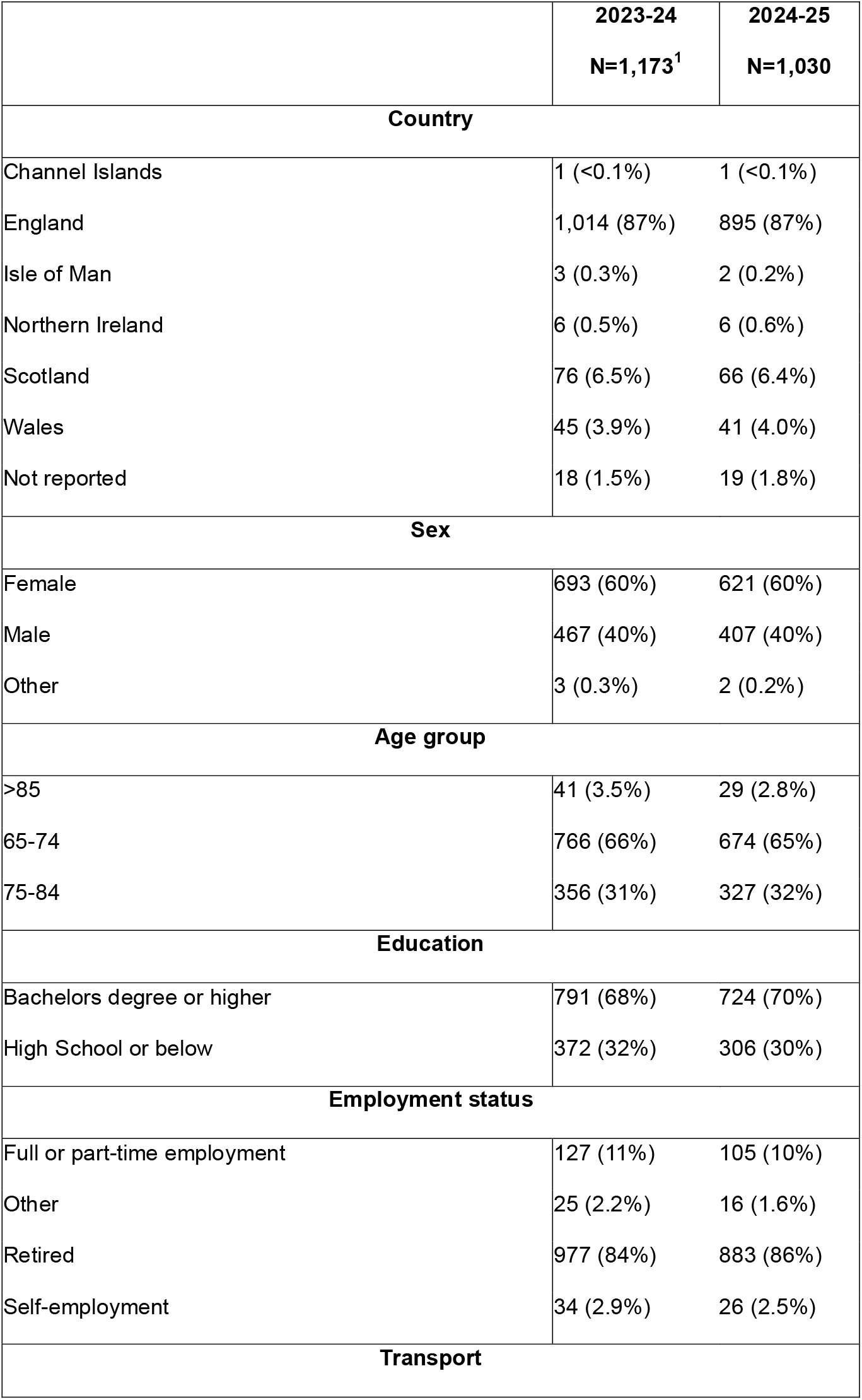
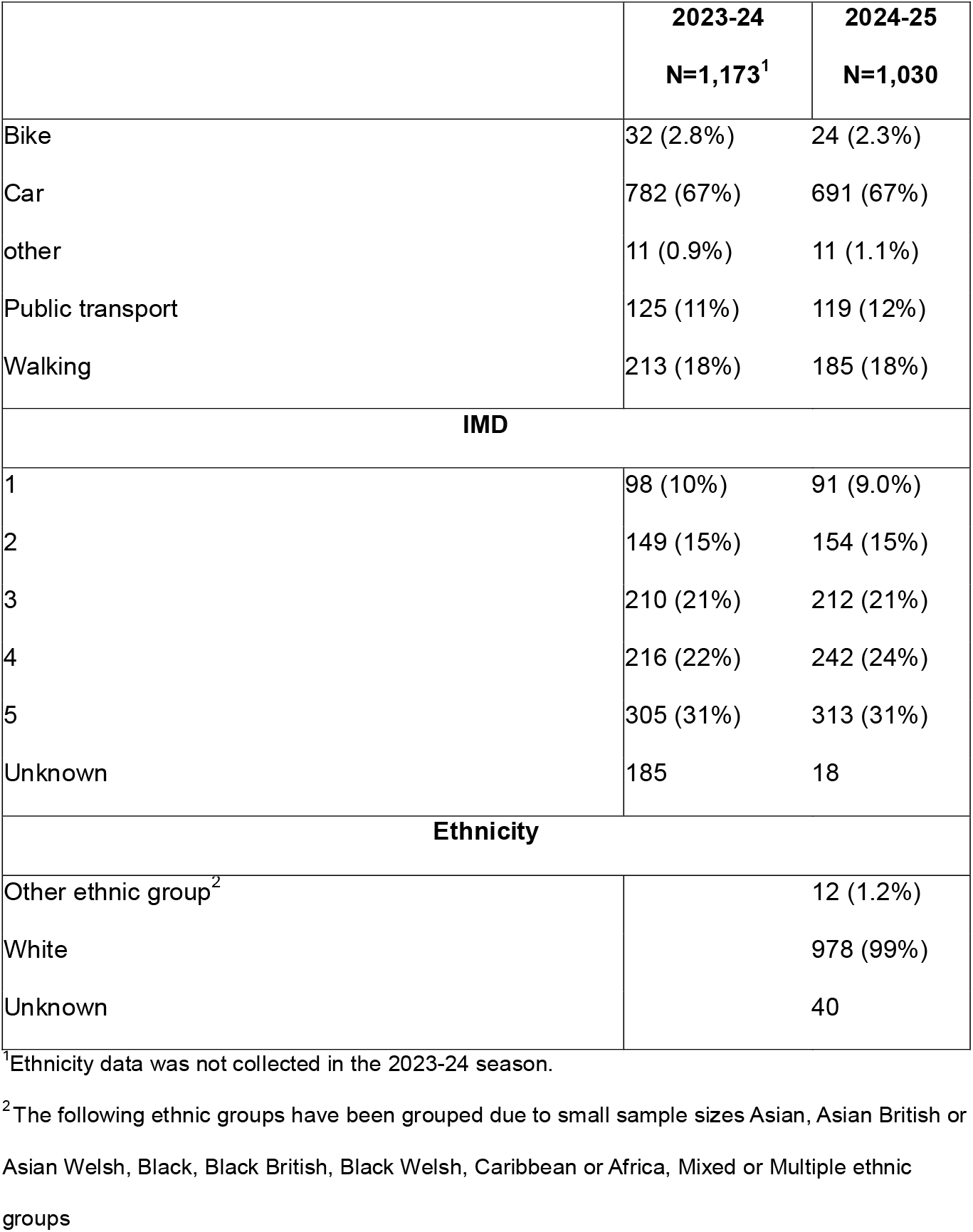
Characteristics of FluSurvey participants from 2023–24 to 2024–25 season who are ≥65.

### S3. Characteristics of vaccinated participants

**Table S3.**
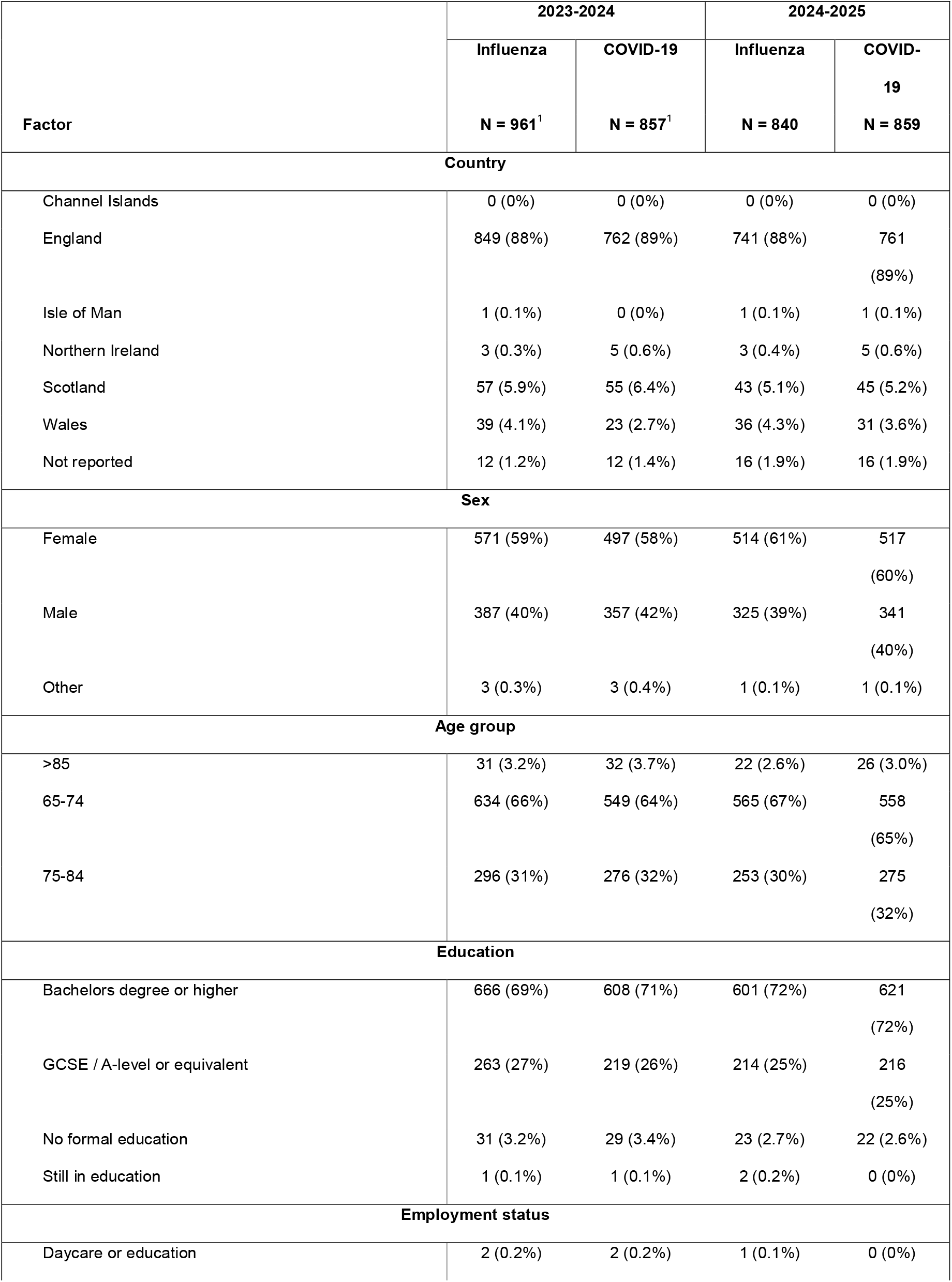
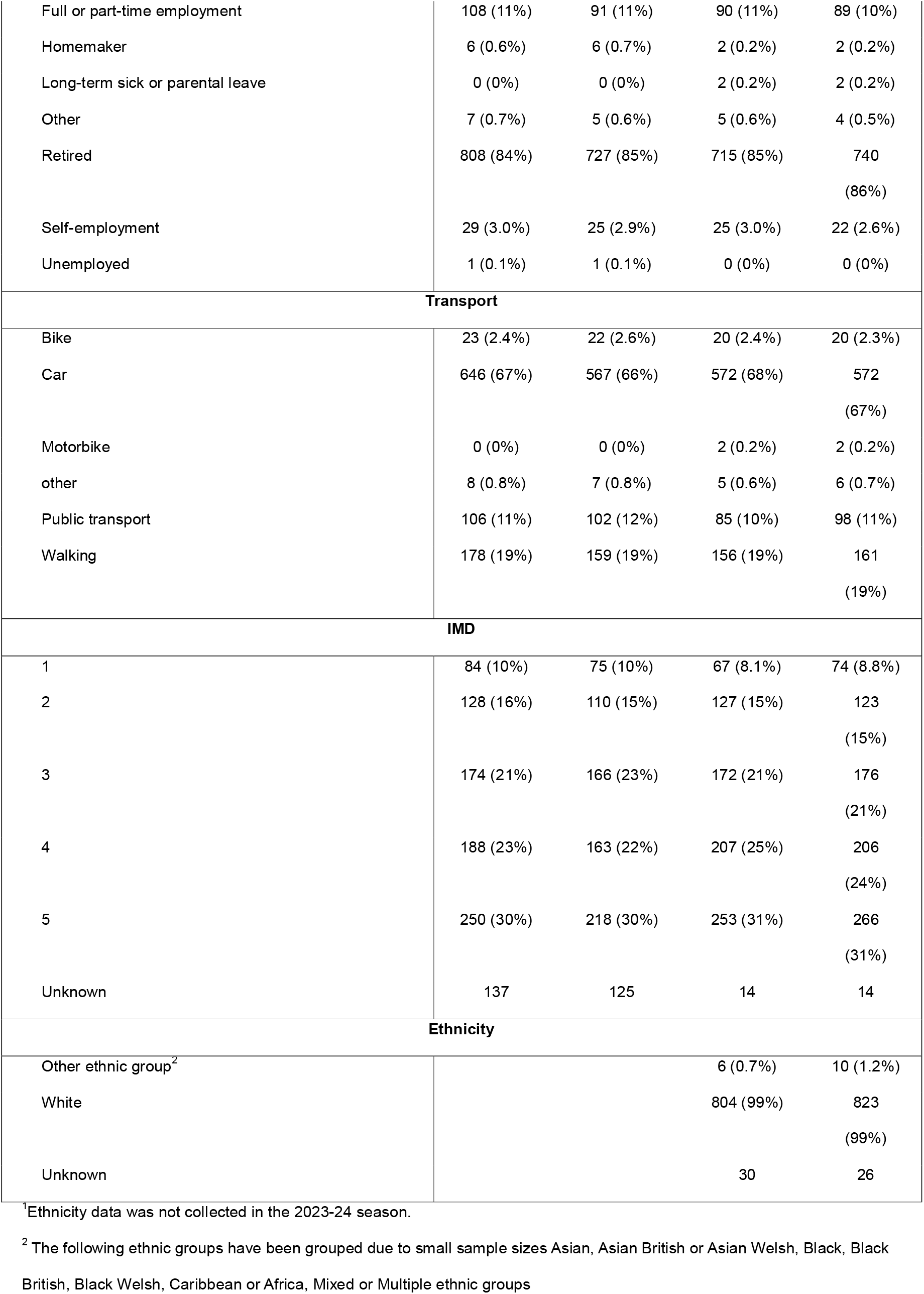
Characteristics of FluSurvey participants vaccinated against influenza and COVID-19 by season and by characteristics (≥65 only)

### S4. Univariable analysis

**Table S4.**
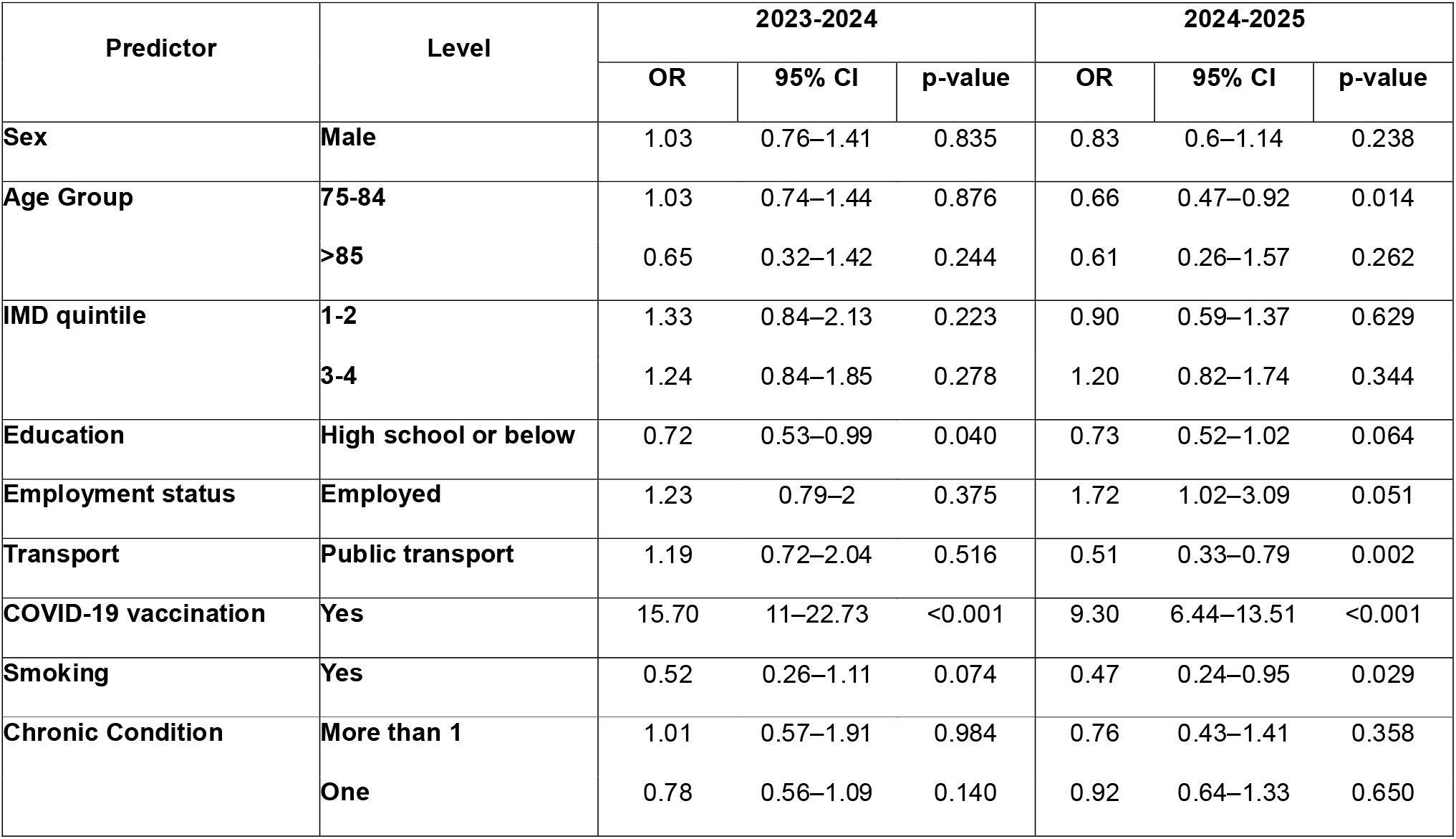
Factors associated with influenza vaccination uptake (age 65++; n = 857 in 2023-2024 and n = 859 in 2024-2025). Unadjusted logistic regression odds ratios with 95% confidence intervals.

**Table S5.**
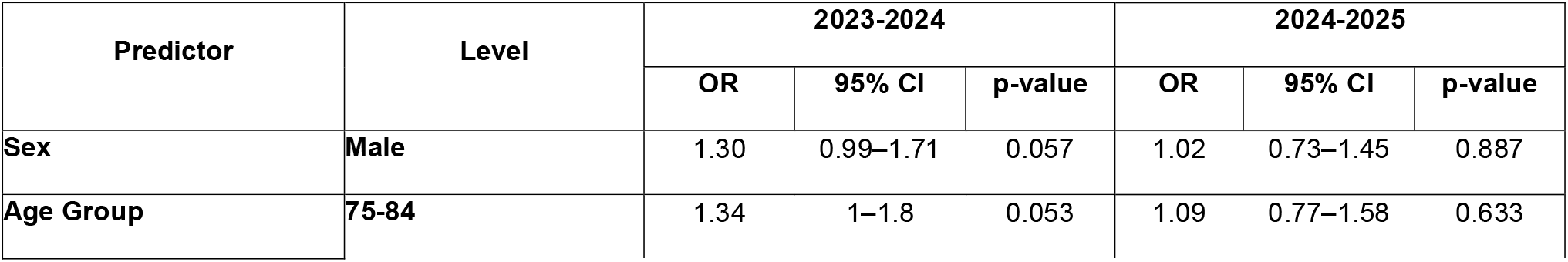
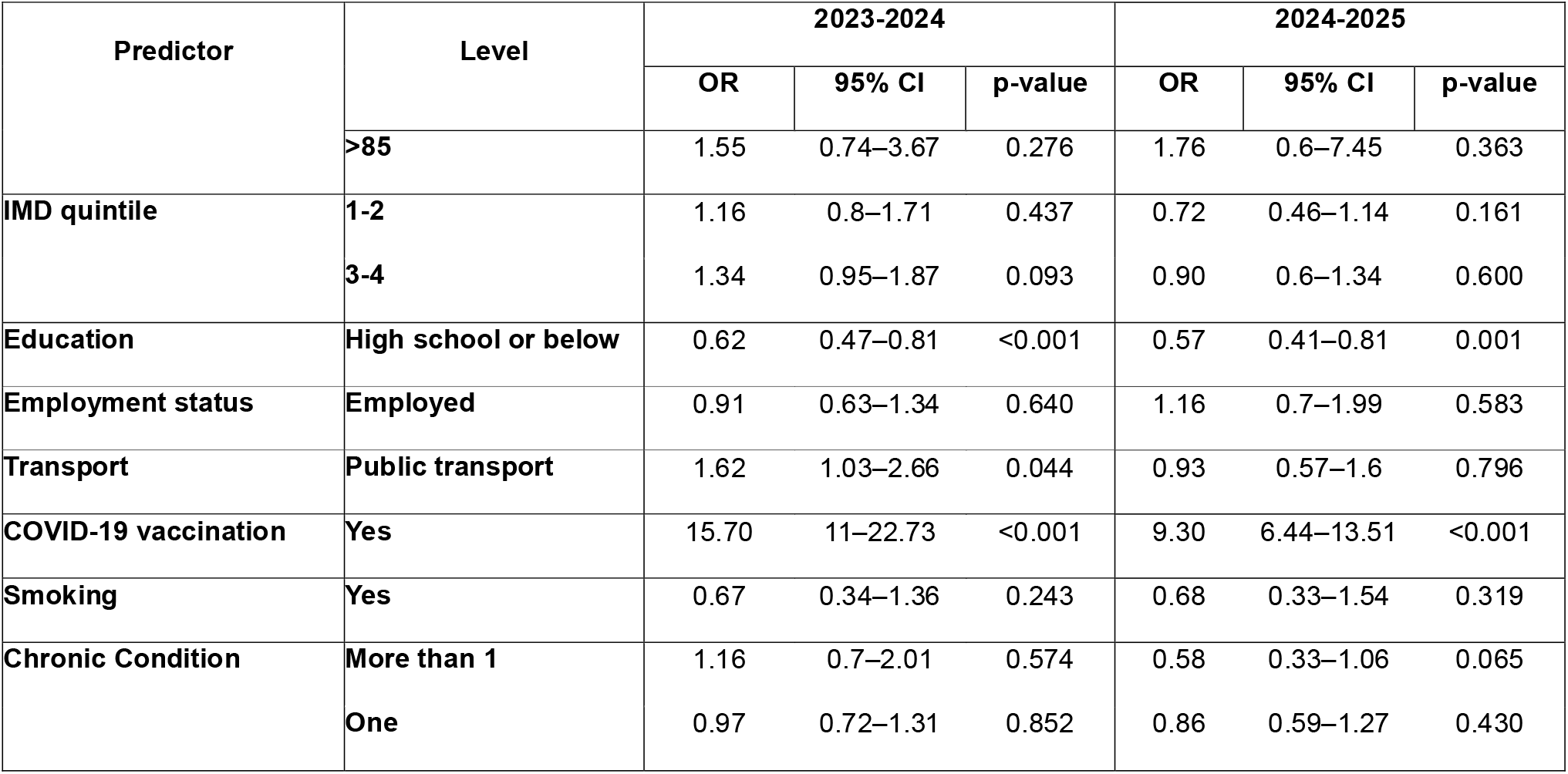
Factors associated with influenza vaccination uptake (age 65++; n = 857 in 2023-2024 and n = 859 in 2024-2025). Unadjusted logistic regression odds ratios with 95% confidence intervals.

