## supplementary material for "Factors associated with vaccination against Seasonal Influenza and SARS-CoV-2 in the UK among FluSurvey participants, 2023-2025"

**S1. Survey phrasing and factor grouping**

Table S1. Survey phrasing for the variables derived in the present analyses are included below. Reference levels are indicated by an Asterix *.

| **Variable** | **Question 2023-24** | **Question 2024-25** | **Notes** |
| --- | --- | --- | --- |
| Country | **What is the first part of your home postcode (the part before the space)?**   - Free text - Don’t know | | *Transformations:*  Postcode mapped to country  Since the first part of each postcode contains multiple areas of varying IMD. Each postcode was mapped to all possible IMD’s and a mean was calculated. |
| Sex | **What is your sex?**   - Male - Female* - Other | | - |
| Age | **What is your date of birth (year and month)?**  Dropdown month selection Dropdown year selection | | *Transformations:*  Age calculated from month & year of birth  Grouped into ≤18, 19-44, 45-64, 65-74*, 75-84 and ≥85 |
| Ethnic group | Not asked | What is your ethnic group?  ONS Census 20b [Ethnic group classifications: Census 2021 - Office for National Statistics](https://www.ons.gov.uk/census/census2021dictionary/variablesbytopic/ethnicgroupnationalidentitylanguageandreligionvariablescensus2021/ethnicgroup/classifications) | *Transformations:*  Remapped to ONS Census 6b [Ethnic group classifications: Census 2021 - Office for National Statistics](https://www.ons.gov.uk/census/census2021dictionary/variablesbytopic/ethnicgroupnationalidentitylanguageandreligionvariablescensus2021/ethnicgroup/classifications) |
| Highest education | **What is the highest level of formal education/qualification that you have?**   - I have no formal qualification - GCSE's, O'levels, CSEs or equivalent - A-levels or equivalent (e.g. Higher, NVQ Level3, BTEC) - Bachelors Degree (BA, BSc) or equivalent - Higher Degree or equivalent (e.g. Masters Degree, PGCE, PhD, Medical Doctorate, Advanced Professional Award) - I am still in education | | *Transformations:* grouped into high school or below and degree or above* |
| Work activity | **What is your work pattern/activity?**   - Paid employment, full time, with at least some work away from home - Paid employment, part time, with at least some work away from home - Working from home (full time) - Working from home (part time) - Self-employed (business owner, farmer, tradesperson, etc.) - Attending daycare/school/college/university - Homemaker (stay at home partner or person with caring responsibilities) - Unemployed - Long-term sick-leave or parental leave - Retired - Other | | *Transformations*: grouped into employed and not employed* |
| Smoking | **Do you smoke tobacco?**   - No* - Yes, occasionally - Yes, daily, fewer than 10 times a day - Yes, daily, 10 or more times a day - I use e-cigarettes only - Dont know/would rather not answer | | *Transformations:*  All “Yes” options combined into “Yes” smoker. |
| Co-morbidities requiring medication | **Do you take regular medication for any of the following medical conditions?**   - No* - Asthma - Diabetes - Chronic lung disorder besides asthma e.g. COPD, emphysema, or other disorders that affect your breathing - Heart disorder - Kidney disorder - An immunocompromising condition (e.g. splenectomy, organ transplant, acquired immune deficiency, cancer treatment) - I would rather not answer | | *Transformations:*  Grouped into either 1 chronic condition or more than 1 chronic condition. |
| Influenza vaccination | **Are you planning to receive a flu vaccine this autumn/winter season?**   - Yes, I'm planning to - Yes, I have got one - No *   I don't know | | - |
| COVID-19 vaccination | **Are you planning to receive a COVID-19 vaccine this season?**   - Yes, I'm planning to - Yes, I've had one - No * - I don't know | | **Are you planning to receive a COVID-19 vaccine this season?**   - Yes, I'm planning to - Yes, I've had one - No, I’m eligible but not planning to - No, I’m not in one of the eligible groups - I don't know |

**S2. Characteristics of participants who are over 65**

Table S2. Characteristics of FluSurvey participants from 2023–24 to 2024–25 season who are ≥65.

|  | **2023-24**  **N=1,173^1^** | **2024-25**  **N=1,030** |
| --- | --- | --- |
| **Country** | | |
| Channel Islands | 1 (<0.1%) | 1 (<0.1%) |
| England | 1,014 (87%) | 895 (87%) |
| Isle of Man | 3 (0.3%) | 2 (0.2%) |
| Northern Ireland | 6 (0.5%) | 6 (0.6%) |
| Scotland | 76 (6.5%) | 66 (6.4%) |
| Wales | 45 (3.9%) | 41 (4.0%) |
| Not reported | 18 (1.5%) | 19 (1.8%) |
| **Sex** | | |
| Female | 693 (60%) | 621 (60%) |
| Male | 467 (40%) | 407 (40%) |
| Other | 3 (0.3%) | 2 (0.2%) |
| **Age group** | | |
| >85 | 41 (3.5%) | 29 (2.8%) |
| 65-74 | 766 (66%) | 674 (65%) |
| 75-84 | 356 (31%) | 327 (32%) |
| **Education** | | |
| Bachelors degree or higher | 791 (68%) | 724 (70%) |
| High School or below | 372 (32%) | 306 (30%) |
| **Employment status** | | |
| Full or part-time employment | 127 (11%) | 105 (10%) |
| Other | 25 (2.2%) | 16 (1.6%) |
| Retired | 977 (84%) | 883 (86%) |
| Self-employment | 34 (2.9%) | 26 (2.5%) |
| **Transport** | | |
| Bike | 32 (2.8%) | 24 (2.3%) |
| Car | 782 (67%) | 691 (67%) |
| other | 11 (0.9%) | 11 (1.1%) |
| Public transport | 125 (11%) | 119 (12%) |
| Walking | 213 (18%) | 185 (18%) |
| **IMD** | | |
| 1 | 98 (10%) | 91 (9.0%) |
| 2 | 149 (15%) | 154 (15%) |
| 3 | 210 (21%) | 212 (21%) |
| 4 | 216 (22%) | 242 (24%) |
| 5 | 305 (31%) | 313 (31%) |
| Unknown | 185 | 18 |
| **Ethnicity** | | |
| Other ethnic group^2^ |  | 12 (1.2%) |
| White |  | 978 (99%) |
| Unknown |  | 40 |
| ^1^Ethnicity data was not collected in the 2023-24 season.  ^2^ The following ethnic groups have been grouped due to small sample sizes Asian, Asian British or Asian Welsh, Black, Black British, Black Welsh, Caribbean or Africa, Mixed or Multiple ethnic groups | | |

**S3. Characteristics of vaccinated participants**

Table S3. Characteristics of FluSurvey participants vaccinated against influenza and COVID-19 by season and by characteristics (≥65 only)

|  | **2023-2024** | | **2024-2025** | |
| --- | --- | --- | --- | --- |
|  | **Influenza** | **COVID-19** | **Influenza** | **COVID-19** |
| **Factor** | **N = 961**^1^ | **N = 857**^1^ | **N = 840** | **N = 859** |
| **Country** | | | | |
| Channel Islands | 0 (0%) | 0 (0%) | 0 (0%) | 0 (0%) |
| England | 849 (88%) | 762 (89%) | 741 (88%) | 761 (89%) |
| Isle of Man | 1 (0.1%) | 0 (0%) | 1 (0.1%) | 1 (0.1%) |
| Northern Ireland | 3 (0.3%) | 5 (0.6%) | 3 (0.4%) | 5 (0.6%) |
| Scotland | 57 (5.9%) | 55 (6.4%) | 43 (5.1%) | 45 (5.2%) |
| Wales | 39 (4.1%) | 23 (2.7%) | 36 (4.3%) | 31 (3.6%) |
| Not reported | 12 (1.2%) | 12 (1.4%) | 16 (1.9%) | 16 (1.9%) |
| **Sex** | | | | |
| Female | 571 (59%) | 497 (58%) | 514 (61%) | 517 (60%) |
| Male | 387 (40%) | 357 (42%) | 325 (39%) | 341 (40%) |
| Other | 3 (0.3%) | 3 (0.4%) | 1 (0.1%) | 1 (0.1%) |
| **Age group** | | | | |
| >85 | 31 (3.2%) | 32 (3.7%) | 22 (2.6%) | 26 (3.0%) |
| 65-74 | 634 (66%) | 549 (64%) | 565 (67%) | 558 (65%) |
| 75-84 | 296 (31%) | 276 (32%) | 253 (30%) | 275 (32%) |
| **Education** | | | | |
| Bachelors degree or higher | 666 (69%) | 608 (71%) | 601 (72%) | 621 (72%) |
| GCSE / A-level or equivalent | 263 (27%) | 219 (26%) | 214 (25%) | 216 (25%) |
| No formal education | 31 (3.2%) | 29 (3.4%) | 23 (2.7%) | 22 (2.6%) |
| Still in education | 1 (0.1%) | 1 (0.1%) | 2 (0.2%) | 0 (0%) |
| **Employment status** | | | | |
| Daycare or education | 2 (0.2%) | 2 (0.2%) | 1 (0.1%) | 0 (0%) |
| Full or part-time employment | 108 (11%) | 91 (11%) | 90 (11%) | 89 (10%) |
| Homemaker | 6 (0.6%) | 6 (0.7%) | 2 (0.2%) | 2 (0.2%) |
| Long-term sick or parental leave | 0 (0%) | 0 (0%) | 2 (0.2%) | 2 (0.2%) |
| Other | 7 (0.7%) | 5 (0.6%) | 5 (0.6%) | 4 (0.5%) |
| Retired | 808 (84%) | 727 (85%) | 715 (85%) | 740 (86%) |
| Self-employment | 29 (3.0%) | 25 (2.9%) | 25 (3.0%) | 22 (2.6%) |
| Unemployed | 1 (0.1%) | 1 (0.1%) | 0 (0%) | 0 (0%) |
| **Transport** | | | | |
| Bike | 23 (2.4%) | 22 (2.6%) | 20 (2.4%) | 20 (2.3%) |
| Car | 646 (67%) | 567 (66%) | 572 (68%) | 572 (67%) |
| Motorbike | 0 (0%) | 0 (0%) | 2 (0.2%) | 2 (0.2%) |
| other | 8 (0.8%) | 7 (0.8%) | 5 (0.6%) | 6 (0.7%) |
| Public transport | 106 (11%) | 102 (12%) | 85 (10%) | 98 (11%) |
| Walking | 178 (19%) | 159 (19%) | 156 (19%) | 161 (19%) |
| **IMD** | | | | |
| 1 | 84 (10%) | 75 (10%) | 67 (8.1%) | 74 (8.8%) |
| 2 | 128 (16%) | 110 (15%) | 127 (15%) | 123 (15%) |
| 3 | 174 (21%) | 166 (23%) | 172 (21%) | 176 (21%) |
| 4 | 188 (23%) | 163 (22%) | 207 (25%) | 206 (24%) |
| 5 | 250 (30%) | 218 (30%) | 253 (31%) | 266 (31%) |
| Unknown | 137 | 125 | 14 | 14 |
| **Ethnicity** | | | | |
| Other ethnic group^2^ |  |  | 6 (0.7%) | 10 (1.2%) |
| White |  |  | 804 (99%) | 823 (99%) |
| Unknown |  |  | 30 | 26 |
| ^1^Ethnicity data was not collected in the 2023-24 season.  ^2^ The following ethnic groups have been grouped due to small sample sizes Asian, Asian British or Asian Welsh, Black, Black British, Black Welsh, Caribbean or Africa, Mixed or Multiple ethnic groups | | | | |

**S4. Univariable analysis**

Table S4. Factors associated with influenza vaccination uptake (age 65++; n = 857 in 2023-2024 and n = 859 in 2024-2025). Unadjusted logistic regression odds ratios with 95% confidence intervals.

| **Predictor** | **Level** | **2023-2024** | | | **2024-2025** | | |
| --- | --- | --- | --- | --- | --- | --- | --- |
|  |  | **OR** | **95% CI** | **p-value** | **OR** | **95% CI** | **p-value** |
| **Sex** | **Male** | 1.03 | 0.76–1.41 | 0.835 | 0.83 | 0.6–1.14 | 0.238 |
| **Age Group** | **75-84** | 1.03 | 0.74–1.44 | 0.876 | 0.66 | 0.47–0.92 | 0.014 |
|  | **>85** | 0.65 | 0.32–1.42 | 0.244 | 0.61 | 0.26–1.57 | 0.262 |
| **IMD quintile** | **1-2** | 1.33 | 0.84–2.13 | 0.223 | 0.90 | 0.59–1.37 | 0.629 |
|  | **3-4** | 1.24 | 0.84–1.85 | 0.278 | 1.20 | 0.82–1.74 | 0.344 |
| **Education** | **High school or below** | 0.72 | 0.53–0.99 | 0.040 | 0.73 | 0.52–1.02 | 0.064 |
| **Employment status** | **Employed** | 1.23 | 0.79–2 | 0.375 | 1.72 | 1.02–3.09 | 0.051 |
| **Transport** | **Public transport** | 1.19 | 0.72–2.04 | 0.516 | 0.51 | 0.33–0.79 | 0.002 |
| **COVID-19 vaccination** | **Yes** | 15.70 | 11–22.73 | <0.001 | 9.30 | 6.44–13.51 | <0.001 |
| **Smoking** | **Yes** | 0.52 | 0.26–1.11 | 0.074 | 0.47 | 0.24–0.95 | 0.029 |
| **Chronic Condition** | **More than 1** | 1.01 | 0.57–1.91 | 0.984 | 0.76 | 0.43–1.41 | 0.358 |
|  | **One** | 0.78 | 0.56–1.09 | 0.140 | 0.92 | 0.64–1.33 | 0.650 |

Table S5. Factors associated with influenza vaccination uptake (age 65++; n = 857 in 2023-2024 and n = 859 in 2024-2025). Unadjusted logistic regression odds ratios with 95% confidence intervals.

| **Predictor** | **Level** | **2023-2024** | | | **2024-2025** | | |
| --- | --- | --- | --- | --- | --- | --- | --- |
|  |  | **OR** | **95% CI** | **p-value** | **OR** | **95% CI** | **p-value** |
| **Sex** | **Male** | 1.30 | 0.99–1.71 | 0.057 | 1.02 | 0.73–1.45 | 0.887 |
| **Age Group** | **75-84** | 1.34 | 1–1.8 | 0.053 | 1.09 | 0.77–1.58 | 0.633 |
|  | **>85** | 1.55 | 0.74–3.67 | 0.276 | 1.76 | 0.6–7.45 | 0.363 |
| **IMD quintile** | **1-2** | 1.16 | 0.8–1.71 | 0.437 | 0.72 | 0.46–1.14 | 0.161 |
|  | **3-4** | 1.34 | 0.95–1.87 | 0.093 | 0.90 | 0.6–1.34 | 0.600 |
| **Education** | **High school or below** | 0.62 | 0.47–0.81 | <0.001 | 0.57 | 0.41–0.81 | 0.001 |
| **Employment status** | **Employed** | 0.91 | 0.63–1.34 | 0.640 | 1.16 | 0.7–1.99 | 0.583 |
| **Transport** | **Public transport** | 1.62 | 1.03–2.66 | 0.044 | 0.93 | 0.57–1.6 | 0.796 |
| **COVID-19 vaccination** | **Yes** | 15.70 | 11–22.73 | <0.001 | 9.30 | 6.44–13.51 | <0.001 |
| **Smoking** | **Yes** | 0.67 | 0.34–1.36 | 0.243 | 0.68 | 0.33–1.54 | 0.319 |
| **Chronic Condition** | **More than 1** | 1.16 | 0.7–2.01 | 0.574 | 0.58 | 0.33–1.06 | 0.065 |
|  | **One** | 0.97 | 0.72–1.31 | 0.852 | 0.86 | 0.59–1.27 | 0.430 |
